# The association of gut microbiome composition with musculoskeletal features in middle-aged and older adults: a two-cohort joint study

**DOI:** 10.1101/2025.02.19.25322523

**Authors:** Ruolin Li, Paul C. Okoro, M. Carola Zillikens, Ramachandran S. Vasan, Shivani Sahni, Fernando Rivadeneira, Douglas P. Kiel, Carolina Medina-Gomez

## Abstract

**Background:** Bones and muscles are connected anatomically, and functionally. Preliminary evidence has shown the gut microbiome influences the aging process of bone and muscle in animal studies. However, such evidence in humans is still scarce. This study aimed to assess the microbiome-bone and microbiome-muscle associations in two cohorts of community-dwelling older adults.

**Methods:** We leveraged information from two large population-based cohorts, i.e., the Rotterdam Study (mean age 62.7 ± 5.6 years; n=1,249) and the Framingham Heart Study (mean age 55.2 ± 9.1 years; n=1,227). For individuals included in this study, gut microbiome 16S rRNA sequencing, musculoskeletal phenotyping derived from DXA images, lifestyle and socioeconomic data, and medication records were available. Per cohort, the 16S rRNA sequencing data, derived from stool, were processed with the DADA2 pipeline and taxonomies were assigned using the SILVA reference database. In addition, the microbiome functional potential was obtained with PICRUSt2. Further, we investigated the association between the human gut microbiome (alpha diversity, genera and predicted functional pathways) and appendicular lean mass (ALM), femoral neck bone mineral density (FN-BMD) and trabecular bone score (TBS) using multilinear regression models controlling for multiple confounders, and performed a joint analysis from both cohorts. Sex-stratified analyses were also conducted.

**Results:** The gut microbiome alpha diversity was not associated with either tested phenotype after accounting for multiple-testing (P>1.67e-02). In the joint analysis, lower abundance of *Oscillibacter* (beta= −.51, 95%CI [−0.74, −.29]), *Anaerotruncus* (beta=-0.41, 95%CI [−0.61, - 0.21]), *Eisenbergiella* (beta=-0.39, 95%CI [−0.59, −.19]) and higher abundance of *Agathobacter* (beta=0.40, 95%CI [0.20, 0.60]) were associated with higher ALM (P<2.0e-04). Lower abundance of *Anaerotruncus* (beta=-0.32, 95%CI [−0.45, −.19]), *Hungatella* (beta=-0.26, 95%CI [−0.38, −.15]) and *Clostridiales bacterium DTU089* (beta=-0.37, 95%CI [−0.55, −.19]) was associated with higher ALM only in females (P< 2.0e-04). Moreover, the *biotin biosynthesis II* pathway was positively associated with ALM (beta=0.44, 95% CI [0.24, 0.64]) (P<1.90e-04) in females while no associations were observed in males. We did not observe any robust association of bone traits with gut microbiome features.

**Conclusion:** Our results indicate that specific genera are associated with ALM in middle-aged and older adults and these associations can present in a sex-specific manner. Overall, our study suggests that the gut microbiome is linked to muscle aging in middle-aged and older adults. However, larger sample sizes are still needed to underpin the specific microbiome features involved.

## 1 Introduction

People worldwide are living longer, and according to the World Health Organization, by 2030, 1 in 6 individuals will be aged 60 years or over.^1^ As the aged population increases, the number of people affected by age-related chronic diseases, such as osteoporosis and sarcopenia, is also growing. Consequently, healthcare services are strained under increasing medical expenses among older adults.

Osteoporosis is a metabolic bone disorder characterized by low bone mass and microarchitectural deterioration of bone tissue, with a consequent increase in bone fragility and susceptibility to fracture.^2^ Bone fractures can result in changes in posture, pain, functional impairment, and the need for surgical repair. Loss of bone mass starts in adulthood after reaching its peak and gradually increases at older ages, especially after menopause.^3^ The regulation of bone mass is a process that includes a complex set of interactions between hormones (parathyroid, gonadal, glucocorticoids), vitamin D, growth factors, and specialized cells related to bone tissue (osteoclasts, osteoblasts, and osteocytes).^4,5^ The maintenance of bone mass depends on the balance between bone resorption and bone formation (bone remodeling). Risk factors for osteoporosis include older age, being female and of European or Asian background, low body mass index (BMI), low level of estrogen or testosterone, smoking, inadequate dietary intake of key nutrients, physical inactivity, lower education level, genetics, and diseases such as hyper(para)thyroidism.^6^

Osteoporosis and sarcopenia often co-occur during the process of musculoskeletal aging. Sarcopenia consists of persistent muscle atrophy characterized by gradual loss of skeletal muscle mass and function (strength).^7^ Older adults with sarcopenia experience increased risk of frailty, disability, injurious falls, hospitalizations, mortality, and reduced quality of life ^8^. Similar to osteoporosis, risk factors for sarcopenia include older age, lower educational level or income, underweight, visceral fat accumulation, low physical activity or protein intake, shorter or longer sleeping time, smoking, cognitive impairment, diabetes mellitus, cardiovascular and respiratory diseases.^9^

The role of the gut microbiome in the development of osteoporosis and sarcopenia has received increased interest given its promising potential in improving musculoskeletal health. The gut microbiome is commonly assessed in the stool and comprises a collection of microorganisms from the digestive tract that impact human physiology through different biological processes. Previous research has indicated that the community of commensal microbes residing in the gut may represent a potentially modifiable factor contributing to muscle and skeletal health.^10-12^ For instance, the gut microbiome can affect the inflammatory environment through effects on the T-cell landscape, which influences osteoclastogenesis and bone loss in mice,^13,14^ and through the production of complex polysaccharides (e.g., short chain fatty acids (SCFA)).^15^ The gut microbiota also interacts with important diet components associated with musculoskeletal health such as vitamin K,^16^ vitamin D,^17^ and calcium.^18,19^ Moreover, the gut microbiota can modulate lipopolysaccharide (LPS) production and various metabolites that directly or indirectly (i.e., through the brain and liver) affect host skeletal muscle metabolism potentially playing a role in sarcopenia etiology.^20^

Despite the growing body of literature supporting the involvement of the gut microbiota in the maintenance of muscle and bone, there are few large clinical investigations in humans of such associations.^21,22,23^ Our study aimed to assess the microbiome-bone and microbiome-muscle associations in two cohorts of community-dwelling older adults, i.e., the Rotterdam Study (RS), and the Framingham Heart Study (FHS).

## 2 Methods

### 2.1 Description of study cohorts and participants

#### 2.1.1 The Rotterdam Study

The Rotterdam Study (RS) is a prospective population-based cohort study established in 1990 to study determinants of disease and disability in Dutch adult individuals.^24^ The RS consists of four sub-cohorts and comprises approximately 18,000 inhabitants of the Ommoord suburb in Rotterdam, predominantly individuals of European background, aged ≥ 40 years.^24^ The fecal collection started in 2012 among the third cohort, the Rotterdam Study III (RS-III). All subjects provided written informed consent before participation in the study. The research was performed following the Declaration of Helsinki. Ethical approval was obtained from the Medical Ethical Committee of Erasmus MC (MEC-02-1015).

#### 2.1.2 The Framingham Study

The Framingham Study (FHS) began in 1948 with the recruitment of a community-based sample of residents from the town of Framingham, Massachusetts, United States and surrounding towns to study the risk factors for heart disease. Over the subsequent decades, the offspring of the original participants were recruited with their spouses into the Offspring Cohort, and finally the children of the Offspring and their spouses were recruited into the third-generation cohort (Gen3). Additional Offspring cohort spouses were also included, and they were evaluated along with the Gen3 participants. Finally, a multi-ethnic cohort, the Omni cohort, was recruited from the same towns as the Gen3 participants and was assessed along with the Gen3 participants. The methods used to collect gut microbiome samples from FHS Gen3, the additional spouses and the Omni cohort participants have been well described in a previous study including their sub-cohort information.^25^ All participants from the FHS provided written informed consent before participation in the study. The research was approved by the Advarra Institutional Review Board.

### 2.2 Stool sample collection, 16S rRNA gene sequencing, and sequence data preprocessing

#### 2.2.1 RS

Details of stool sample collection for RS can be found elsewhere.^26^ Briefly, RS participants sent stool samples collected at home to Erasmus MC Rotterdam through regular mail, and once in-site samples were stored at −20°C. Participants also filled in a short questionnaire including the date and time of defecation and current or recent antibiotic use (past year). An automated stool DNA isolation kit (Diasorin, Saluggia, Italy) was used to isolate bacterial DNA from the stool. The V3 and V4 hypervariable regions of the bacterial 16S rRNA gene were then amplified and sequenced on an Illumina MiSeq platform.

#### 2.2.2 FHS

Stool samples were collected in 100% ethanol, and bacterial DNA was extracted using a Qiagen custom protocol and stored at −20°C. The V4 hypervariable region of the 16S rRNA gene was targeted and amplified with the 515F primer and unique reverse barcode primers from the Golay primer set.^27^ The amplified PCR products were sequenced on an Illumina MiSeq machine with the 2 x 150 base pair paired-end protocol. Detailed information can be found in a previous publication.^21^

### 2.3 Preprocessing of 16S rRNA samples in the FHS and RS

For each study cohort, the resulting 16S rRNA gene amplicon sequencing data were separately processed with DADA2 ^28^ denoising pipeline. Briefly, the paired-end sequence data were de-multiplexed. Then DADA2 denoising, filtering, trimming, merging, chimera removal, and taxonomic assignment were executed in R, next, an amplicon sequence variants (ASVs) table was obtained as output. For RS, a naive Bayesian classifier from the Ribosomal Database Project (RDP 2.12), and SILVA 16S database release 138.1 were used to reconstruct the taxonomic composition of the studied communities.^29,30^ ASV filtering was applied, i.e., each ASV had to comprise at least 0.5% of the total reads and be present in at least 1% of the study samples to remain in the dataset.^31^ Taxonomical classification from kingdom through genus was assigned if the bootstrap confidence score was above 50 (i.e., binning posterior probability cut-off was 0.5). Species assignment was only reported if an ASV could be matched to a species with a confidence score of 100 binning posterior probability. For FHS, resulting ASVs were assigned taxonomies using the SILVA reference database release 132 for the genus-level classification, and version 128 was used for the species-level classification. For downstream analysis, Phyloseq ^32^ version 1.38.0 was used to generate two separate taxa count tables from the ASVs count tables; one glommed at the genus level and another at the species level.

### 2.4 Metagenome prediction

We used PICRUSt2 ^33^ to predict MetaCyc functional pathways ^34^ abundance from the 16S rRNA amplicon sequence data. Briefly, PICRUSt2 predicts pathway abundances based on the number of taxa contributing to each particular pathway based on a filtered ASV-table as described in section 2.3. In total, 360 pathways were predicted in RS and 394 in FHS. Then, we used the “findCorrelation” function from the caret R package ^35^ to remove features that were highly correlated (Pearson correlation > 0.9) with each other. In cases where two features are highly correlated, the function considers the mean absolute correlation of each feature with all other ASVs present in the data and removes the one with the highest mean absolute correlation. Association tests were performed in a set of 91 functional features in RS and 112 in FHS.

### 2.5 Skeletal and muscle measurements

#### 2.5.1 RS

DXA scans from the total body, lumbar spine and femoral neck were obtained using an iDXA densitometer (GE Lunar Corp, Madison, WI, USA). A trained technician performed and verified all bone scans. Scans were then analyzed using enCORE (v 13.60). TBS was calculated using the TBS iNsight software (version 4, Med-Imaps, Geneva, Switzerland)^36^ from DXA images of the lumbar spine, adjusting for tissue thickness derived from the same image. Briefly, the TBS is a gray-level texture measurement that captures the lumbar trabecular component and its characteristics.^37^ A higher score indicates better microarchitecture which leads to stronger bones with increased fracture resistance.

Lean body mass measurements were obtained from the total body scans. Appendicular lean mass (ALM) was calculated as the sum of the lean tissue from the arms and legs. In addition, maximum handgrip strength (HGS) was measured by a single trained examinator using a hydraulic hand dynamometer (Fabrication Enterprises Inc., White Plains, NY, USA). Maximum HGS was computed as the maximum value (kg) out of three trials performed with the non-dominant hand.

#### 2.5.2 FHS

BMD was measured at Gen3 Exam 2 (2008-2011) at the hip and spine (average BMD of L2-L4) using a GE Lunar Prodigy fan-beam densitometer (GE Healthcare, Inc.), using standard positioning recommended by the manufacturer. BMD was measured in grams per centimeter squared (g/cm^2^). The right hip was scanned unless there was a history of previous fracture or hip replacement, in which case the left side was scanned. Participants were not scanned if there were concerns about radiation or the possibility of pregnancy. Participants weighing 136 kg or more did not undergo the assessment. TBS was calculated from acquired DXA scans of the spine (i.e., L2-L4), accounting for soft tissue thickness, using the TBS iNsight® software (version 4, Medimaps Group, Geneva, Switzerland) as previously described.^38^

Following the recommendations of the International Society for Clinical Densitometry, a reliability assessment of the Prodigy machine was conducted. Twenty volunteers were measured three times by the same technician at both the hip and spine, with repositioning after each scan. The percent coefficients of variation (%CV) for the Prodigy device were 1.8, 2.3, 1.2 and 1.1 for the femoral neck, trochanter, total hip and L2-4 regions, respectively. Further information regarding this study can be found in a previous publication.^39^

Lean mass measurements were calculated from the whole-body scans collected at Exam 2 (2008-2011) employing the same densitometer, using standard positioning recommended by the manufacturer and applying GE Healthcare software Inc. Encore v16. In some cases, the participant was too large to include all the extremities, whereupon the individual was positioned in the center to measure visceral adipose tissue, causing the arms to be outside of the scan limits. In other instances, even the visceral adipose area was too large for the scan field. In those cases, the individual was positioned with a complete half of the whole-body image within the scan field, and the opposite side of the body was derived from the side that was included using the hemiscan feature. During the cleaning process scans were removed when limbs were outside of the scan field, and also when the difference in lean mass or fat mass between arms or legs differed by more than three standard deviations (sd) of the mean. ALM was computed by adding lean mass from both arms and legs.

### 2.6 Covariates

#### 2.6.1 RS

For every participant, height and weight were measured at the research center, and BMI was calculated as weight (kg) divided by height (m)^2^. Total body fat mass was derived from the DXA scans and divided by weight to calculate fat percent. Type 2 diabetes (T2D) status was determined as follows: fasting glucose > 6.9 mmol/L, non-fasting glucose >11.0 mmol/L, or the use of antidiabetic medication including insulin in the RS cohort. In addition, two study physicians independently determined all potential T2D events. In case of disagreement, consensus was sought from an endocrinologist.^40^ Information on smoking status was obtained through a questionnaire filled in at the same visit and participants were classified as current or non-current smokers. Information on diet was obtained about five years before stool sampling. A 389-item food frequency questionnaire (FFQ) ^41^ was used to determine dietary intake. Moreover, a diet quality score was generated as the sum of individual components each measuring the participants’ adherence (yes/no) to 14 items of the Dutch dietary guidelines. A higher score reflects better diet quality.^41^ Medication use (i.e., metformin, proton pump inhibitors (PPI) and lipid-lowering medications) was retrieved from general practitioners, pharmacies’ databases, nationwide medical registries, or follow-up examinations.

Important technical covariates can affect the microbiome profiles and must be considered in any statistical analysis. Based on previous publications with the data,^31^ we included sampling season, TimeInMail (i.e., the days between the deposition and the storage of the sample in the research center), DNA isolation batch and sequencing batch (i.e., whether a sample was re-sequenced in an attempt to increase the sequence quality).^31^

#### 2.6.2 FHS

Race was self-reported by participants as one of four categories: African American, Asian, Hispanic, White or Other. Participants with T2D were defined as those currently using a medication indicated for the treatment of diabetes or having either a fasting blood glucose level of ≥ 126 mg/dL or a random blood glucose level of ≥ 200 mg/dL. Participants with normal blood glucose < 126 mg/dL and taking no diabetes medications were considered non-diabetic. Hospitalization was dichotomized as whether the participant had a hospitalization in the past year or not. A diet quality index (DQI) was created using the information in the Willett 126-item semi-quantitative FFQs that were collected in 2008-2011.^42^ This FFQ has been validated in previous studies.^42,43^ Smoking status was based on self-report, and individuals were classified as current smokers vs non-smokers (past smokers and those who had never smoked).

### 2.7 Statistical analysis

Descriptive characteristics of the participants of each cohort were compared using Student’s t-tests for continuous normally distributed variables or Wilcoxon signed-rank tests otherwise. To compare dichotomous variables chi-squared tests were applied.

To assess the potential relationship between the gut microbiome and the musculoskeletal outcomes (i.e., FN-BMD, TBS and ALM), we performed analyses at three different levels, i.e., the microbiome alpha diversity (as assessed by the number of observed genera and the Shannon index at the genus level), the genera differential abundance, and the functional potential, estimated by the MetaCyc pathways. For the RS cohort, the covariates included in the FN-BMD and TBS models were age, sex, BMI (kg/m^2^), T2D (yes/no), current smoking (yes/no), diet quality score, PPI use (yes/no), metformin use (yes/no), use of lipid-lowering medications (yes/no), sequencing batch, DNA isolation batch, time in mail, and sampling season. Whereas for the FHS, age, sex, race (self-reported), BMI, T2D (yes/no), hospitalizations in the past year (yes/no), current smoking (yes/no), DQI, ^44,45^ total number of medications used (as count), PPI use (yes/no), and metformin use (yes/no). For the ALM analyses, in both cohorts, BMI was replaced by height and body fat percent.

All models were performed in all individuals, and males and females separately because of well-known differences in bone and muscle characteristics between sexes. Alpha diversity analyses were performed using multivariable linear regression models with each musculoskeletal outcome used as a dependent variable. The differential abundance analyses for genera and pathways were run in each cohort independently with MaAsLin2 (v 1.8.0),^46^ using the musculoskeletal traits as independent variables and applying the default parameters (min_abundance = 0.0, min_prevalence = 0.1, analysis_method= “LM”, normalization= “TSS”, transform= “LOG”, max_significance= 0.25). The Benjamini & Hochberg method was used for multiple-testing correction.

Only genera and pathways that overlapped across the two cohorts were included in the joint analyses. These joint analyses were performed using two different approaches. First, we combined the effect sizes weighted by their standard errors, using a fixed-effects or a random-effects model in *metafor.*^47^ The choice of model was based on whether there was significant heterogeneity of effect sizes across the two cohorts. A random-effects model was used for features presenting a heterogeneity P < 0.05; otherwise, a fixed-effects model was used. In addition, we also performed the meta-analysis by combining P-values following a sample size weighted-Fisher approach, as implemented in the *metapro* R package.^48^ We applied multiple-testing correction as follows, alpha diversity significance threshold 1.67e-02 (i.e., 0.05/3 phenotypes) and differential abundance significance threshold (0.05/ (number of tested genera or pathways x number of tested phenotypes). Therefore, for the taxa analyses, the significance threshold was 2.0e-4 while for the pathways analyses, it was 1.90e-04.

Associations between taxa or pathways that were associated with ALM, were also tested for association with muscle strength (measured as maximum HGS) in RS, using a multivariable linear model with centered-log-ratio (CLR) transformed taxa abundance as the independent variable while adjusting for the same covariates used for the ALM analyses.

## 3. Results

### 3.1 Characteristics of study populations and participants

Our study included participants from two large observational studies: the Rotterdam Study (RS), and the Framingham Heart Study (FHS). Stool samples received from each participant (RS=1,420, FHS=1,424) were processed for 16S rRNA amplicon sequencing. In RS, after excluding samples from participants taking antibiotics within 30 days prior to stool collection, or with missing data or samples that stayed at room temperature for more than seven days, 1,249 samples remained for analysis. In FHS, after excluding samples from participants with antibiotic use within 30 days before collection or colon surgery within one year before stool collection, or participants with missing covariates, 1,227 samples remained. A figure describing the inclusion and exclusion details for each cohort can be found in **Supplementary Material, Figure S1**.

The average age of RS participants was 62.7 years (range: 52.4-93.2 years), while FHS participants were younger with an average age of 55.2 years (range: 32.0-89.0 years) (**Table 1A**). Compared to FHS participants, RS participants had lower BMI (P <0.001), higher body fat percent (P <0.001), higher ALM (P <0.001), lower femoral neck BMD (FN-BMD, P <0.001), and lower average TBS (P <0.001) (**Table 1A**). Similar differences were also observed in the sex-specific comparisons across the two cohorts (**Table 1B**).

**Table 1A.** Characteristics of the participants in this study. This study included participants from two large observational studies: the Rotterdam Study (RS), and the Framingham Heart Study (FHS) with gut microbiome valid samples.

|  | RS (N=1,249) | FHS (N=1,227) | P |
| --- | --- | --- | --- |
| Age (y; mean ± sd) | 62.66 ± 5.65 | 55.18 ± 9.06 | <0.001* |
| Sex (men, %) | 41.71 (521) | 44.34 (544) | 0.20 |
| Height (m; mean ± sd) | 1.71 ± 0.09 | 1.69 ± 0.09 | <0.001* |
| BMI (kg/m<sup>2</sup>; mean ± sd) | 27.29 ± 4.31 | 27.97 ± 5.48 | <0.001* |
| Body fat percentage (%; mean ± sd)<br>(n=1,249 RS; n=1,105 FHS) | 35.32 ± 7.30 | 32.59 ± 9.12 | <0.001* |
| Prevalence of type 2 diabetes (%) | 11.93 (149) | 8.72 (107) | 0.11 |
| Appendicular lean mass (kg; mean ± sd)<br>(n=1,249 RS; n=1,105 FHS) | 22.66 ± 5.20 | 21.24 ± 5.53 | <0.001* |
| Maximum handgrip strength (kg; mean ± sd) | 31.06 ± 10.33 | NA | NA |
| <sup>1</sup>Femoral neck bone mineral density (g/cm<sup>2</sup>; mean ± sd) (n=1,240 RS; n=1,197 FHS) | 0.92 ± 0.14 | 0.96 ± 0.14 | <0.001* |
| Average trabecular bone score (mean ± sd)<br>(n=1,248 RS; n=991 FHS) | 1.36 ± 0.08 | 1.43 ± 0.08 | <0.001* |
| Stool microbiome alpha diversity-Shannon index | 2.96 ± 0.36 | 2.49 ± 0.59 | <0.001* |

“NA”: Data not available. “\*”: Significant P value.
<sup>1</sup> Femoral neck bone mineral density and appendicular lean mass were measured by specific iDXA densitometer in RS cohort, and measured by a GE Lunar Prodigy fan beam densitometer (GE Healthcare, Inc.) in FHS cohort.

**Table 1B.** Characteristics of the participants stratified by sex. This study included participants from two large observational studies: the Rotterdam Study (RS), and the Framingham Heart Study (FHS) with gut microbiome valid samples.

|  | Male |  |  | Female |  |  |
| --- | --- | --- | --- | --- | --- | --- |
|  | RS (N=521) | FHS (N=544) | P | RS (N=728) | FHS (N=683) | P |
| Age (y; mean ± sd) | 62.65 ± 5.72 | 55.62 ± 9.21 | <0.001* | 62.67 ± 5.60 | 54.82 ± 8.93 | <0.001* |
| Height (m; mean ± sd) | 1.79 ± 0.07 | 1.77 ± 0.07 | <0.001* | 1.65 ± 0.06 | 1.63 ± 0.06 | <0.001* |
| BMI (kg/m<sup>2</sup>; mean ± sd) | 27.48 ± 3.91 | 28.97 ± 4.76 | <0.001* | 27.15 ± 4.58 | 27.18 ± 5.88 | 0.90 |
| Body fat percentage (%; mean ± sd)<br>(male: n=521 RS; n=462 FHS<br>female: n=728 RS; n=643 FHS) | 30.42 ± 5.75 | 26.87 ± 6.67 | <0.001* | 38.82 ± 6.20 | 36.70 ± 8.40 | <0.001* |
| Prevalence of type 2 diabetes (%) | 17.66% (92) | 11.95% (65) | 0.011* | 7.83% (57) | 6.15% (42) | 0.26 |
| Appendicular lean mass (kg; mean ± sd)<br>(male: n=521 RS; n=462 FHS<br>female: n=728 RS; n=643 FHS) | 27.68 ± 3.52 | 26.79 ± 3.43 | <0.001* | 19.06 ± 2.56 | 17.25 ± 2.46 | <0.001* |
| Maximum handgrip strength (kg; mean ± sd) | 40.17 ± 8.86 | NA | NA | 24.54 ± 4.98 | NA | NA |
| Femoral neck bone mineral density (g/cm<sup>2</sup>; mean ± sd)<br>(male: n=520 RS; n=530 FHS<br>female: n=720 RS; n=667 FHS) | 0.97 ± 0.13 | 1.01 ± 0.13 | <0.001* | 0.88 ± 0.12 | 0.93 ± 0.13 | <0.001* |
| Average trabecular bone score (mean ± sd)<br>(male: n=521 RS; n=441 FHS<br>female: n=727 RS; n=550 FHS) | 1.39 ± 0.08 | 1.44 ± 0.08 | <0.001* | 1.34 ± 0.08 | 1.42 ± 0.09 | <0.001* |
| Stool microbiome alpha diversity-Shannon index | 2.96 ± 0.35 | 2.46± 0.59 | <0.001* | 2.95 ± 0.37 | 2.52 ± 0.58 | <0.001* |

“NA”: Data not available. “\*”: Significant P-value.

### 3.2 Association between alpha diversity and musculoskeletal phenotypes

The microbiome alpha diversity was not associated with any of the musculoskeletal phenotypes using the stringent multiple-testing threshold (P< 1.67e-02) (**Supplementary Table S1**). Yet, the Shannon index showed a positive association with FN-BMD in RS at a nominal level (P< 0.05) (**Table 2**). No significant results were observed in the sex-stratified analyses.

**Table 2.** Association between Shannon diversity index and musculoskeletal phenotypes. This study included participants from two large observational studies: the Rotterdam Study (RS), and the Framingham Heart Study (FHS) with gut microbiome valid samples.

| Outcome | Joint-analysis |  |  | Rotterdam Study |  |  | Framingham Heart Study |  |  |
| --- | --- | --- | --- | --- | --- | --- | --- | --- | --- |
|  | Shannon index<br>(95% CI) | <sup>1</sup>P. FE | <sup>2</sup>P. Fisher (direction) | Shannon index<br>(95% CI) | P | N | Shannon index<br>(95% CI) | P | N |
| Appendicular lean mass | -0.04<br>(-0.09, 0.02) | 0.20 | 0.39 (-) | -0.02<br>(-0.41, 0.37) | 0.93 | 1249 | -0.04<br>(-0.09, 0.02) | 0.20 | 1105 |
| Femoral neck bone<br>mineral density | 0.02<br>(1.40e-03, 0.04) | 0.04 | 0.08 (+) | 0.02<br>(1.57e-03, 0.04) | 0.03 | 1249 | 8.00e-03<br>(-0.08, 0.20) | 0.85 | 1197 |
| Trabecular bone score | 2.70e-03<br>(-9.70e-03, 0.02) | 0.67 | 0.95 (+) | 2.94e-03<br>(-9.53e-03, 0.02) | 0.64 | 1249 | -8.00e-03<br>(-0.11, 0.09) | 0.87 | 991 |

<sup>1</sup>: Results obtained by fixed effect (FE) meta-analysis
<sup>2</sup>: Results obtained by P value combination following a sample size weighted-Fisher approach.

### 3.3 Association between microbial taxa and musculoskeletal phenotypes

Assessing all available data, four genera were associated (P<2e-04) with ALM, i.e., *Oscillibacter* (beta= −.51, 95%CI [−0.74, −.29], N_not0_=1,737), *Anaerotruncus* (beta=-0.41, 95%CI [−0.61, −.21], N_not0_=984), *Agathobacter* (beta=0.40, 95%CI [0.20, 0.60], N_not0_=2,249), and *Eisenbergiella* (beta=-0.39, 95%CI [−0.59, −.19], N_not0_=667) (**Table 3A)**. with three of the four genera negatively associated with ALM and one genus positively associated with ALM. These genera showed no significant effect heterogeneity across the studied cohorts. Moreover, the results for *Oscillibacter* and *Anaerotruncus* remained significant using the weighted-Fisher approach. No genus was associated with FN-BMD or TBS after multiple-testing correction (P>2.0e-04). Among the four genera associated with ALM in the analysis including all individuals, higher *Eisenbergiella* abundance was associated with lower HGS (beta=-0.37, 95% CI [−0.65, −.08], P=0.01) in RS. Results can be found in **Supplementary Table S6.**

**Table 3A.** Genera associated with ALM. This study included participants from two large observational studies: the Rotterdam Study (RS), and the Framingham Heart Study (FHS) with gut microbiome valid samples.

| Genera | Joint-analysis |  |  | Rotterdam Study |  |  | Framingham Heart Study |  |  |
| --- | --- | --- | --- | --- | --- | --- | --- | --- | --- |
|  | Beta<br>(95% CI) | <sup>1</sup>P.FE | <sup>2</sup>P.Fisher<br>(direction) | Beta<br>(95% CI) | <sup>3</sup>P.adjusted | N<sub>not0</sub> | Beta<br>(95% CI) | <sup>3</sup>P.adjusted | N<sub>not0</sub> |
| Oscillibacter | -0.51<br>(-0.74, -0.29) | 9.35e-06 | 1.94e-5 (-) | -0.62<br>(-0.94, -0.30) | 5.84e-03 | 690 | -0.41<br>(-0.73, -0.08) | 0.13 | 1047 |
| Anaerotruncus | -0.41<br>(-0.61, -0.21) | 4.80e-05 | 8.31 e-5 (-) | -0.35<br>(-0.58, -0.13) | 0.04 | 241 | -0.61<br>(-1.02, -0.20) | 0.06 | 743 |
| Agathobacter | 0.40<br>(0.20, 0.60) | 8.77e-05 | 2.10e-4 (+) | 0.40<br>(0.16, 0.64) | 0.03 | 1192 | 0.40<br>(0.03, 0.77) | 0.20 | 1057 |
| Eisenbergiella | -0.39<br>(-0.59, -0.19) | 1.12e-04 | 3.95e-4 (-) | -0.37<br>(-0.59, -0.15) | 0.02 | 175 | -0.49<br>(-0.97, -0.02) | 0.22 | 492 |

<sup>1</sup>: Results obtained by the combination of effect sizes meta-analysis implemented in *metafor* package. P.FE < 2.0e-04 is the cut-off value for significance after multiple- testing correction. FE = fixed effects model.
<sup>2</sup> Results obtained by P value combination following a sample size weighted-Fisher approach. P.Fisher < 2.0e-04 is the cut-off value for significance after multiple-testing correction.
<sup>3</sup>: Results obtained using the Benjamini & Hochberg correction method based on the original P value.

In the sex-stratified analysis, we found three genera associated with ALM in females, *Anaerotruncus* (beta=-0.32, 95%CI [−0.45, −.19], N_not0_=618), *Hungatella* (beta=-0.26, 95%CI [−0.38, −.15], N_not0_=402), and *Clostridiales bacterium DTU089* (beta = −.37, 95%CI [−0.55, −.19], N_not0_=624) (**Table 3B)**; while showing no effect in males (Table 3B). To note, in analyses of RS males, *Hungatella* was filtered out due to low prevalence (N=49, 9.4%). We did not observe any significant (P>2.0e-04) associations between the interrogated genera with either FN-BMD or TBS in the sex-stratified analyses. The ten genera showing the highest evidence of association with each musculoskeletal trait can be found in **Supplementary Table S2A-S2C.** Mainly the direction of effect was consistent across cohorts for these genera. In RS females, the ALM-associated *Anaerotruncus* was also associated with lower HGS (beta=-0.41, 95% CI [−0.75, −.06], P=0.02). Results can be found in **Supplementary Table S6.**

**Table 3B.** Genera associated with ALM in sex-stratified analyses. This study included participants from two large observational studies: the Rotterdam Study (RS), and the Framingham Heart Study (FHS) with gut microbiome valid samples.

|  |  | Joint-analysis |  |  | Rotterdam Study |  |  | Framingham Heart Study |  |  |
| --- | --- | --- | --- | --- | --- | --- | --- | --- | --- | --- |
| Genera | Subgroup | Beta<br>(95% CI) | <sup>1</sup>P.FE | <sup>2</sup>P.Fisher<br>(direction) | Beta<br>(95% CI) | <sup>3</sup>P.adjusted | N<sub>not0</sub> | Beta<br>(95% CI) | <sup>3</sup>P.adjusted | N<sub>not0</sub> |
| Anaerotruncus | Female | -0.32<br>(-0.45, -0.19) | 1.50e-06 | 2.90e-06 (-) | -0.28<br>(-0.42, -0.14) | 0.01 | 156 | -0.49<br>(-0.80, -0.19) | 0.11 | 462 |
|  | Male | -0.12<br>(-0.28, 0.04) | 0.14 | 0.16 (-) | -0.09<br>(-0.27, 0.10) | 0.81 | 85 | -0.21<br>(-0.51, 0.09) | 0.64 | 281 |
| Hungatella | Female | -0.26<br>(-0.38, -0.15) | 1.14e-05 | 7.97e-05 (-) | -0.26<br>(-0.38, -0.13) | 0.01 | 85 | -0.31<br>(-0.63, 0.01) | 0.34 | 317 |
|  | Male | NA | NA | NA | NA | NA | NA | -0.13<br>(-0.46, 0.19) | 0.42 | 177 |
| Clostridiales<br>bacterium<br>DTU089 | Female | -0.37<br>(-0.55, -0.19) | 7.38e-05 | 1.34e-04 (-) | -0.33<br>(-0.54, -0.12) | 0.08 | 462 | -0.48<br>(-0.84, -0.12) | 0.17 | 320 |
|  | Male | -0.15<br>(-0.35, 0.06) | 0.16 | 0.24 (-) | -0.14<br>(-0.40, 0.11) | 0.71 | 205 | -0.16<br>(-0.52, 0.20) | 0.83 | 175 |

<sup>1</sup>: Results obtained by the combination of effect sizes meta-analysis implemented in *metafor* package. P.FE < 2.0 e-04 is the cut-off value for significance after multiple-testing correction. FE = fixed effects model.
<sup>2</sup>: Results obtained by P value combination following a sample size weighted-Fisher approach. P.Fisher < 2.0 e-04 is the cut-off value for significance after multiple-testing correction.
<sup>3</sup>: Results obtained using the Benjamini & Hochberg correction method based on original p values.
“NA”: Data not available (taxon was filtered out due to low prevalence).

We also assessed the associations of gut microbiome with each musculoskeletal phenotype within each study cohort. In RS, we identified 13 genera associated with ALM, one genus associated with FN-BMD and one genus associated with TBS (all P.adjusted < 0.1). For the sex-stratified analyses, in males, one genus was associated with ALM and another one with TBS (P.adjusted < 0.1), whereas in females, five genera were associated with ALM, two genera with FN-BMD and one other genus with TBS. In FHS, we identified 16 genera associated with ALM, four associated with FN-BMD, and three with TBS (all P.adjusted < 0.1). For the sex-stratified analysis, in men, one genus was associated with FN-BMD (P.adjusted < 0.1), whereas in women, two genera were associated with ALM, and one genus associated with FN-BMD. Results can be found in **Supplementary Tables S3A-S3C**.

### 3.4 Association between microbial functional pathways and musculoskeletal phenotypes

We also tested the association of predicted functional pathways and musculoskeletal phenotypes. Only in females, we observed a significant association (P<1.90e-04). Higher abundance of the *biotin biosynthesis II* pathway was associated with higher ALM (beta=0.44, 95% CI [0.24, 0.64], P= 1.55e-05, N_not0_ =1,281). This pathway showed no association with ALM in males (beta=0.05, 95% CI [−0.15, 0.25], P= 0.62, N_not0_ = 915). Analyses performed with the sample size weighted-Fisher approach reached consistent findings. Yet, the *biotin biosynthesis II* pathway was not associated with HGS in female RS participants (beta= 0.08, 95% CI [−0.11, 0.26], P=0.42). The top 10 associated pathways for each musculoskeletal phenotype can be found in **Supplementary Table S4A-S4C,** in most cases, the direction of effect was consistent across cohorts. Cohort specific associations (P.adjusted <0.1) can be found in **Supplementary Table S5A-S5C**.

## Discussion

Musculoskeletal health is an important component of healthy aging. The gut microbiome has been implicated in the metabolism of muscle and bone. In our study, we tested the association between microbiome profiles and musculoskeletal phenotypes in two large cohorts (RS and FHS). Our study highlighted four microbial genera associated with ALM in the meta-analysis of both study cohorts; we also identified three genera and one functional pathway associated with ALM in females. Two of the identified genera additionally showed a directionally consistent association with hand grip strength. In contrast, we did not find any significant association, after multiple-test correction, of microbiome features with FN-BMD or TBS.

Microbiome studies and especially meta-analyses are confronted with many challenges. The differences in study protocols are particularly important when analyzing data from more than one cohort. In this study, the harmonization of the analytical pipeline, data collection and sample profiling were carried out differently in RS and FHS. Many researchers have assessed the impact of DNA extraction,^49,50^ sequenced variable region,^50-52^ and other technical variables, for example, sample storage time and temperature ^53^ in the generation of microbiome profiles. These and other confounding variables in the experimental design hinder the comparison of studies and contribute to the lack of replicability common in microbiome research.^54^ In a previous meta-analysis, combining results of the association of microbiome profiles and high-resolution peripheral quantitative computed tomography parameters from FHS and the Osteoporotic Fractures in Men Study (MrOS) participants, the authors estimated that providing the observed effect sizes, studies will need at least 6,000 individuals to have sufficient power to evidence the association of microbiome composition with skeletal traits.^21^ Similarly, our study also lacks sufficient power to unveil such associations. In contrast, we discovered genera associated with lean mass mirroring studies describing microbiome features associated with lean mass in the healthy population at lower sample sizes.^22,23^

To account for the potential heterogeneity present in our study, we followed different strategies. A standard meta-analysis approach, which relies on the exposure effect size. Also, we implemented another strategy contingent on a combination of probability tests (P-values) using a weighted-Fisher method, in which only the sample size and direction of the effect are considered, and as such is robust to “batch effects” at the expense of statistical power.^55^ Given our limited statistical power, we report cohort-specific association summary statistics at a lax significance threshold in the supplementary material.

Altogether, we found sufficient evidence for the association of ALM with four gut microbiota genera. *Anaerotruncus* was negatively associated with ALM across the set of applied tests in the combined analyses, but also in the female-specific analyses. *Anaerotruncus* is a butyrate-producing genus ^56^ and, as such, it is considered part of a healthy microbiome, associated with muscle aging amelioration in mice after yeast protein supplementation.^57^ Nevertheless, an increased abundance of this genus has been associated with the intake of a high-fat or Western diet,^58,59^ and the development of obesity and other pro-inflammatory diseases in healthy adults with such a diet.^59^ These results highlight the complex nature of the human gut microbiota, where the effect of a specific taxon can be context-dependent, being interspecific interactions with other bacteria and diet important factors conditioning its contribution to human health. Other bacteria showing a consistent negative association with ALM are *Oscillibacter* and *Eisenbergiella*. *Oscillibacter spp.* were significantly diminished in obese adults in a small observational study (N=26, 17 obese participants) ^60^ and it has also been associated with muscle aging amelioration in mice.^57^ A recent paper,^61^ characterized the *Oscillobacter* cholesterol-metabolizing capabilities of at least three human gut *Oscillobacter* isolates. This, coupled with its diet responsiveness and its involvement in the maintenance of gut barrier integrity, ^62^ makes it an interesting genus to study. However, following the tree of life classification, more than thirty species are branching from this genus. ^63^ It is then, important to note that the resolution of 16S rRNA sequencing studies is a major limitation as species and even strains belonging to the same group can have dissimilar effects on human health. On the other hand, *Eisenbergiella,* a gram-negative, non-motile, non-spore-producing bacteria was previously associated with the ratio of ALM to body weight in Japanese women from the general population (N=495, average age 50.8 (sd: 12.8) years old).^64^ On the contrary, our results show an association between lower levels of *Eisenbergiella* and higher ALM and HGS. Although the data acquisition in both studies differed, Sugimura *et al* used bioelectrical impedance analysis (BIA), the most likely explanation for the contrasting findings can be difference in age, ethnic background, geographic location and dietary intake of the participants, or the normalization strategy and confounders used in the association models. *Agathobacter,* which was positively associated with ALM in the present study, has been associated with higher grip strength and gait speed in a study that involved 276 community-dwelling Chinese older women.^65^

Body composition differs between men and women. Women have proportionally more fat mass and men have more muscle mass. Aging reduces the level of testosterone in women and estrogen in men. The pattern of hormonal decline differs by sex, with women showing a precipitous loss of estrogen during menopause and men losing testosterone gradually throughout life, starting in the third decade. Moreover, the loss of sex hormones is associated with the age-related decline in bone and skeletal muscle mass.^66^ Consequently, we analyzed the association between gut microbiome and musculoskeletal traits in a sex-specific manner. Besides *Anaerotruncus*, we identified two other genera *Hungatella* and *Clostridiales bacterium DTU089* negatively associated with ALM in females. *Hungatella* is a genus comprising at least three species. Recently one of these species, *Hungatella effluvia,* was associated with sarcopenia severity in a shot-gun metagenomics analysis of samples from community-based Chinese adults (N=1,417).^67^ Nevertheless, it is noteworthy to mention this genus was not analyzed in RS males because of low prevalence (i.e., excluded by taxa filtering). On the other hand, *Clostridiales bacterium DTU089* was previously associated with lower tibia cortical volumetric BMD in a study comprising FHS and MrOS.^21^ Furthermore, in this latter cohort, *Clostridiales bacterium DTU089* was present in lower abundance in men with lower protein intake.^68^ Lower BMD and lower protein intake are associated with worse muscle health and therefore consistent with the findings of our study. Aside from microbial genera, our study identified the *biotin biosynthesis II* pathway as positively associated with ALM in females. Biotin, a water-soluble vitamin used as a cofactor of enzymes involved in carboxylation reactions, plays an essential role in maintaining metabolic homeostasis,^69^ and boosts protein biosynthesis through amino acid catabolism.^70^ Previous evidence has shown that biotin deficiency reduces body weight at the expense of muscular mass.^71^ Yet, both biotin excess and biotin deficiency alter skeletal muscle metabolism.^72^ While the effects of biotin on women’s reproductive system have not been established, in mice, both the deficiency and the excess of biotin are associated with an increased estradiol serum concentration.^73^ Even though biotin has also been positively associated with testosterone production in male mice,^74^ we did not find an association of the biotin pathway with ALM in men, possibly because the testosterone decline follows a moderate trajectory in men and thus a higher statistical power is needed. Despite these interesting observations, we did not have sufficient statistical power for instance to explore the effect of menopause in the female-specific associations.

Despite the different lines of evidence from pre-clinical and clinical studies depicting a convincing role of the gut microbiota in bone metabolism,^10^ we did not find any robust association between skeletal traits and the gut microbiome. The most likely explanation is that we are still underpowered to find these associations in observational cross-sectional studies.^75^ Moreover, the largest study on muscle mass and gut microbiome to date, showed that the species *Coprococcus comes*, *Dorea longicatena*, and *Eubacterium ventriosum* were associated with higher muscle mass and BMD at the femoral neck and total hip.^23^ Nevertheless, this study used a targeted approach in which only 50 prevalent and well-characterized microbiota species of the human gut microbiome were surveyed. Therefore, we emphasize the urgent need for collaboration across multiple cohorts to increase the statistical power to study gut microbiome-bone associations in the future. Besides, the development of strategies to integrate shotgun and 16s sequencing data will allow data harmonization from multiple sources, ^76^ presenting a great opportunity to enhance robust studies in the microbiome field.

Several characteristics of our study are worth noting. Compared to previous studies, our study has several strengths. Firstly, both cohorts are deeply phenotyped, which enabled us to account for the influence of important confounders. Secondly, we applied two types of joint analyses that rely on different assumptions to warrant the robustness of our findings. Our study also has limitations that need to be acknowledged. Firstly, we are confined to the 16S rRNA sequencing technology, which provides limited power to investigate taxa at a higher resolution, for example, at the species level, and prevents the direct assessment of microbial genes and pathways. Secondly, our study used a cross-sectional design and thus could not determine whether the identified associations are causal or not. Thirdly, the data of muscle strength was not available in both cohorts, making the relevant findings with regards to muscle strength lacking validation. Future studies are warranted to investigate the mechanisms underlying the effects of the gut microbiome in the musculoskeletal system metabolism or vice versa. Alternatively, animal experiments in combination with stool microbiota transplantation or interventional studies are now on the horizon and would provide a better understanding of this matter.

## Conclusion

To conclude, our study identified several consistent associations between the gut microbiome and muscle health in middle-aged and older adults, suggesting there is likely a gut-muscle link through the musculoskeletal aging process in old adulthood. With the future availability of shotgun sequencing, it is possible to gain additional insights with regard to the specific species that influence muscle mass and strength maintenance. This study provides evidence that targeting the gut microbiota and their associated enzymatic and functional capacities might be an effective approach to improve muscle health in middle-aged and older adults.

## Data Availability

All data produced in the present study are available upon reasonable request to the authors.

## Acknowledgements

The generation and management of the 16S microbiome data for the Generation R Study and the Rotterdam Study was executed by the Human Genotyping Facility of the Genetic Laboratory of the Department of Internal Medicine, Erasmus MC, University Medical Center Rotterdam, the Netherlands. We thank Nahid El Faquir and Jolande Verkroost-Van Heemst for their help in sample collection and registration. We thank Kamal Arabe, Hedayat Razawy, Karan Singh Asra, Pelle van der Wal, Sergio Chavez and Djawad Radjabzadeh for their help in DNA isolation and sequencing, and Joost Verlouw, Dr. Constanza Vallerga and Marijn Verkerk for their help with the bioinformatic analyses. We thank Dr. Cindy Boer, Dr. Robert Kraaij and Prof. Joyce van Meurs for overseeing the quality control of the generated datasets.

The Rotterdam Study gratefully acknowledge the contribution of the inhabitants, general practitioners, and pharmacists of the Ommoord district. The Rotterdam Study is funded by Erasmus Medical Center and Erasmus University, Rotterdam, Netherlands Organization for the Health Research and Development (ZonMw), the Research Institute for Diseases in the Elderly (RIDE), the Ministry of Education, Culture and Science, the Ministry for Health, Welfare and Sports, the European Commission (DG XII), and the Municipality of Rotterdam.

Douglas P. Kiel and Shivani Sahni’s effort, and a portion of the gut microbiome genotyping, were supported by the National Institute of Arthritis and Musculoskeletal and Skin Diseases R01 AR061445. The work was also funded by the National Heart, Lung and Blood Institute’s Framingham Heart Study (FHS [contract numbers HHS N268201500001I, 75N92019D00031 and N01-HC 25195]). The sequencing of the FHS 16S rRNA gene was supported by grant number R01 HL131015. DXA scans in the FHS were supported by grant number R01 AR041398 and the TBS data acquisition was supported by a grant from Dairy Management Inc.

