## Supplementary Tables and Supplementary Figure legend for "The association of gut microbiome composition with musculoskeletal features in middle-aged and older adults: a two-cohort joint study" (from another version of this preprint: version 1, 2025-02-21)

**Table S1. Association between alpha diversity and musculoskeletal phenotypes.** The joint analysis included participants from two large observational studies: the Rotterdam Study (RS), and the Framingham Heart Study (FHS) with gut microbiome valid samples.

| | | Joint analysis* | | | Rotterdam Study | | | Framingham Study | | |
|---|---|---|---|---|---|---|---|---|---|---|
| | | Effect $\beta$ (95%CI) | P. FE** | P. Fisher (direction)# | Effect $\beta$ (95%CI) | P* | Nnot0 | Effect $\beta$ (95%CI) | P* | Nnot0 |
| Overall | **Appendicular lean mass** | | | | | | | | | |
| | Shannon | -0.04 (-0.09, 0.02) | 0.20 | 0.39 (-) | -0.02 (-0.41, 0.37) | 0.93 | 1249 | -0.04 (-0.09, 0.02) | 0.20 | 1105 |
| | Richness | 1.20e-03 (-1.00e-04, 2.60e-03) | 0.07 | 0.24 (+) | -3.77e-03 (-0.01, 6.93e-03) | 0.49 | 1249 | 1.31e-03 (-6.27e-05, 2.69e-03) | 0.06 | 1105 |
| | **Femoral neck bone mineral density** | | | | | | | | | |
| | Shannon | 0.02 (1.40e-03, 0.04) | 0.04 | 0.08 (+) | 0.02 (1.57e-03, 0.04) | 0.03 | 1249 | 8.00e-03 (-0.08, 0.10) | 0.85 | 1197 |
| | Richness | 2.00e-04 (-3.00e-04, 7.00e-04) | 0.40 | 0.31 (+) | 8.80e-05 (-4.45e-04, 6.21e-04) | 0.75 | 1249 | 2.00e-03 (-1.00e-03, 4.00e-03) | 0.19 | 1197 |
| | **Trabecular bone score** | | | | | | | | | |
| | Shannon | 2.70e-03 (-9.70e-03, 0.02) | 0.67 | 0.95 (+) | 2.94e-03 (-9.53e-03, 0.02) | 0.64 | 1249 | -8.00e-03 (-0.11, 0.09) | 0.87 | 991 |
| | Richness | 1.00e-04 (-2.00e-04, 4.00e-04) | 0.52 | 0.76 (+) | 1.15e-04 (-2.28e-04, 4.57e-04) | 0.51 | 1249 | 0.00 (-3.00e-03, 3.00e-03) | 1 | 991 |
| | **Appendicular lean mass** | | | | | | | | | |
| | Shannon | -0.02 (-0.10, 0.06) | 0.57 | 0.58 (-) | -0.23 | 0.56 | 521 | -0.02 | 0.60 | 462 |

| | | | | | | | | | |
|---|---|---|---|---|---|---|---|---|---|
| Male | | | | | (-0.98, 0.53) | | | (-0.10, 0.06) | | |
| | Richness | 1.70e-03 (-8.00e-04, 4.20e-03) | 0.19 | 0.50 (+) | -9.79e-03 (-0.03, 0.01) | 0.33 | 521 | 1.92e-03 (-7.24e-04, 4.55e-03) | 0.15 | 462 |
| | **Femoral neck bone mineral density** | | | | | | | | |
| | Shannon | 0.03 (-6.50e-03, 0.06) | 0.12 | 0.25 (+) | 0.03 (-5.31e-03, 0.06) | 0.1 | 521 | -9.00e-03 (-0.14, 0.12) | 0.89 | 530 |
| | Richness | -1.00e-04 (-9.00e-04, 8.00e-04) | 0.89 | 1.00 (+) | -1.16e-04 (-1.00e-03, 7.68e-04) | 0.80 | 521 | 1.00e-03 (-3.00e-03, 5.00e-03) | 0.65 | 530 |
| | **Trabecular bone score** | | | | | | | | |
| | Shannon | 0.01 (-4.80e-03, 0.03) | 0.14 | 0.33 (+) | 0.02 (-3.92e-03, 0.04) | 0.11 | 521 | -0.06 (-0.20, 0.08) | 0.41 | 441 |
| | Richness | 0.00 (-5.00e-04, 6.00e-04) | 0.86 | 1.00 (-) | 6.63e-05 (-4.66e-04, 5.98e-04) | 0.81 | 521 | -1.00e-3 (-6.00e-03, 4.00e-03) | 0.73 | 441 |
| Female | **Appendicular lean mass** | | | | | | | | |
| | Shannon | 0.04 (-8.70e-03, 0.08) | 0.11 | 0.18 (+) | 0.13 (-0.28, 0.54) | 0.54 | 728 | 0.04 (-0.01, 0.08) | 0.13 | 643 |
| | Richness | 1.00e-03 (-5.00e-04, 2.40e-03) | 0.19 | 0.40 (+) | -7.28e-05 (-0.01, 0.01) | 0.99 | 728 | 9.76e-04 (-4.90e-04, 2.44e-03) | 0.19 | 643 |
| | **Femoral neck bone mineral density** | | | | | | | | |
| | Shannon | 0.02 (-6.40e-03, 0.04) | 0.16 | 0.20 (+) | 0.01 (-8.88e-03, 0.04) | 0.22 | 728 | 0.05 (-0.06, 0.16) | 0.37 | 667 |
| | Richness | 3.00e-04 (-3.00e-04, 1.00e-03) | 0.30 | 0.19 (+) | 3.02e-04 (-3.61e-04, 9.65e-04) | 0.37 | 728 | 2.00e-03 (-1.00e-03, 6.00e-03) | 0.20 | 667 |
| | **Trabecular bone score** | | | | | | | | |
| | Shannon | -3.80e-03 (-0.02, 0.01) | 0.63 | 1.00 (+) | -4.53e-03 (-0.02, 0.01) | 0.58 | 728 | 0.05 (-0.09, 0.19) | 0.50 | 550 |
| | Richness | 2.00e-04 | 0.48 | 0.56 (+) | 1.51e-04 | 0.51 | 728 | 1.00e-03 | 0.63 | 550 |

| | | | | | | | | |
|---|---|---|---|---|---|---|---|---|
| | | (-3.00e-04, 6.00e-04) | | | (-2.98e-04, 5.99e-04) | | | (-3.00e-03, 6.00e-03) |

*P<0.017 is significance cutoff after multiple testing correction

**P value obtained from meta-analysis with fixed effects (FE) model

### P value obtained from meta-analysis by P value combination following sample size weighted-Fisher approach

**Table S2A. Top 10 appendicular lean mass associated taxa; overall and sex stratified**. The joint analysis included participants from two large observational studies: the Rotterdam Study (RS), and the Framingham Heart Study (FHS) with gut microbiome valid samples.

| | | Joint analysis* | | | Rotterdam Study | | | Framingham Study | | |
|---|---|---|---|---|---|---|---|---|---|---|
| | **Taxa** | **Effect $\beta$ (95%CI)** | **P. FE**** | **P. Fisher (direction)#** | **Effect $\beta$ (95%CI)** | **P.adjusted** | **Nnot0** | **Effect $\beta$ (95%CI)** | **P.adjusted** | **Nnot0** |
| Overall | Oscillibacter | -0.51 (-0.74, -0.29) | 9.35e-06 | 1.94e-05 (-) | -0.62 (-0.94 , -0.30) | 5.84e-03 | 690 | -0.41 (-0.73, -0.08) | 0.13 | 1047 |
| | Anaerotruncus | -0.41 (-0.61, -0.21) | 4.80e-05 | 8.31e-05 (-) | -0.35 (-0.58 , -0.13) | 0.04 | 241 | -0.61 (-1.02, -0.20) | 0.06 | 743 |
| | Agathobacter | 0.40 (0.20, 0.60) | 8.77e-05 | 2.10e-04 (+) | 0.40 (0.16 , 0.64) | 0.02 | 1192 | 0.40 (0.03, 0.77) | 0.20 | 1057 |
| | Eisenbergiella | -0.39 (-0.59, -0.19) | 1.12e-04 | 3.95e-04 (-) | -0.37 (-0.59 , -0.15) | 0.02 | 175 | -0.49 (-0.97, -0.02) | 0.22 | 492 |
| | DTU089 | -0.42 (-0.67, -0.18) | 6.26e-04 | 1.13e-03 (-) | -0.38 (-0.66, -0.10) | 0.09 | 509 | -0.54 (-1.02, -0.06) | 0.18 | 495 |
| | Erysipelatoclostridium | -0.47 (-0.75, -0.20) | 7.64e-04 | 1.77e-03 (-) | -0.45 (-0.79, -0.12) | 0.09 | 467 | -0.51 (-0.98, -0.03) | 0.21 | 756 |
| | Hungatella | -0.3 (-0.47, -0.12) | 1.11e-03 | 3.38e-03 (-) | -0.27 (-0.47 , -0.08) | 0.07 | 134 | -0.42 (-0.86, 0.03) | 0.29 | 494 |
| | Romboutsia | 0.53 (0.19, 0.86) | 2.00e-03 | 1.06e-03 (+) | 0.27 (-0.16, 0.70) | 0.57 | 963 | 0.92 (0.39, 1.45) | 0.02 | 664 |
| | CAG.56 | 0.45 (0.16, 0.75) | 2.42e-03 | 2.63e-03 (+) | 0.34 (-0.01, 0.69) | 0.27 | 666 | 0.71 (0.18, 1.24) | 0.09 | 574 |
| | Clostridium.sensu.stricto.1 | 0.56 (0.20, 0.93) | 2.68e-03 | 5.46e-03 (+) | 0.57 (0.05, 1.10) | 0.20 | 891 | 0.55 (0.04, 1.07) | 0.21 | 532 |
| Male | Oscillibacter | -0.31 (-0.48, -0.13) | 5.43e-04 | 2.96e-04 (-) | -0.52 (-0.79 , -0.24) | 0.02 | 267 | -0.17 (-0.39, 0.06) | 0.62 | 432 |
| | Howardella | 0.23 (0.08, 0.38) | 2.13e-03 | 4.98e-03 (+) | 0.20 (-0.02, 0.43) | 0.48 | 143 | 0.25 (0.06, 0.44) | 0.26 | 86 |
| | Alistipes | -0.31 (-0.51, -0.11) | 2.30e-03 | 2.73e-03 (-) | -0.45 (-0.77 , -0.13) | 0.14 | 467 | -0.22 (-0.47, 0.03) | 0.5 | 450 |
| | Allisonella | 0.15 (0.03, 0.27) | 0.01 | 0.01 (+) | 0.11 (-0.03, 0.25) | 0.55 | 56 | 0.25 (0.02, 0.48) | 0.37 | 64 |

| | | | | | | | | | | |
|---|---|---|---|---|---|---|---|---|---|---|
| | Holdemanella | 0.38<br>(0.08, 0.68) | 0.01 | 0.02<br>(+) | 0.44<br>(-2.01e-03 , 0.88) | 0.39 | 228 | 0.33<br>(-0.08, 0.74) | 0.56 | 118 |
| | Eisenbergiella | -0.16<br>(-0.30, -0.03) | 0.02 | 0.06<br>(-) | -0.17<br>(-0.32 , -0.03) | 0.27 | 64 | -0.11<br>(-0.45, 0.24) | 0.88 | 153 |
| | Dorea | 0.16<br>(0.01, 0.32) | 0.03 | 0.06<br>(+) | 0.16<br>(-0.01, 0.33) | 0.45 | 501 | 0.17<br>(-0.14, 0.48) | 0.76 | 426 |
| | Erysipelatoclostridium | -0.23<br>(-0.45, -4.54e-03) | 0.05 | 0.08<br>(-) | -0.25<br>(-0.54, 0.03) | 0.50 | 185 | -0.19<br>(-0.53, 0.16) | 0.77 | 298 |
| | Roseburia | 0.16<br>(2.43e-03, 0.31) | 0.05 | 0.07<br>(+) | 0.14<br>(-0.05, 0.32) | 0.62 | 504 | 0.2<br>(-0.08, 0.48) | 0.63 | 447 |
| | Clostridium.sensu.stricto.1 | 0.32<br>(-8.06e-04, 0.63) | 0.05 | 0.09<br>(+) | 0.30<br>(-0.17, 0.77) | 0.67 | 370 | 0.33<br>(-0.10, 0.76) | 0.59 | 253 |
| Female | Anaerotruncus | -0.32<br>(-0.45, -0.19) | 1.50e-06 | 2.90e-06 (-) | -0.28<br>(-0.42 , -0.14) | 0.01 | 156 | -0.49<br>(-0.80, -0.19) | 0.11 | 462 |
| | Hungatella | -0.26<br>(-0.38, -0.15) | 1.14e-05 | 7.97e-05 (-) | -0.26<br>(-0.38 , -0.13) | 8.04e-03 | 85 | -0.31<br>(-0.63, 0.01) | 0.34 | 317 |
| | DTU089 | -0.37<br>(-0.55, -0.19) | 7.38e-05 | 1.34e-04 (-) | -0.33<br>(-0.54 , -0.12) | 0.08 | 304 | -0.48<br>(-0.84, -0.12) | 0.17 | 320 |
| | Faecalibacterium | 0.28<br>(0.12, 0.44) | 5.46e-04 | 6.37e-04<br>(+) | 0.22<br>(0.03, 0.41) | 0.28 | 712 | 0.41<br>(0.13, 0.69) | 0.13 | 613 |
| | Prevotella_9 | 0.52<br>(0.22, 0.81) | 5.69e-04 | 9.99e-04<br>(+) | 0.63<br>(0.18 , 1.07) | 0.14 | 246 | 0.43<br>(0.04, 0.82) | 0.28 | 140 |
| | Agathobacter | 0.27<br>(0.11, 0.42) | 6.53e-04 | 1.41e-03<br>(+) | 0.27<br>(0.07 , 0.46) | 0.15 | 689 | 0.27<br>(0.02, 0.52) | 0.29 | 608 |
| | Eisenbergiella | -0.26<br>(-0.42, -0.10) | 1.34e-03 | 1.59e-03 (-) | -0.21<br>(-0.39, -0.03) | 0.28 | 111 | -0.44<br>(-0.77, -0.10) | 0.17 | 339 |
| | UBA1819 | -0.27<br>(-0.45, -0.10) | 1.88e-03 | 2.04e-03 (-) | -0.20<br>(-0.42, 0.02) | 0.46 | 471 | -0.40<br>(-0.68, -0.12) | 0.15 | 536 |
| | Erysipelatoclostridium | -0.31<br>(-0.52, -0.11) | 2.87e-03 | 5.95e-03 (-) | -0.29<br>(-0.55 , -0.04) | 0.28 | 282 | -0.35<br>(-0.69, 2.88e-03) | 0.33 | 458 |

| | | | | | | | | | | |
|---|---|---|---|---|---|---|---|---|---|---|
| | Romboutsia | 0.36<br>(0.11, 0.61) | 5.11e-03 | 4.02e-03<br>(+) | 0.17<br>(-0.17, 0.50) | 0.79 | 560 | 0.6<br>(0.22, 0.97) | 0.11 | 363 |

*P<0.0002 is significance cutoff after multiple testing correction

**P value obtained from meta-analysis with fixed effects (FE) model

### P value obtained from meta-analysis by P value combination following sample size weighted-Fisher approach

**Table S2B. Top 10 femoral neck bone mineral density associated taxa; overall and sex stratified**. The joint analysis included participants from two large observational studies: the Rotterdam Study (RS), and the Framingham Heart Study (FHS) with gut microbiome valid samples.

| | | Joint analysis* | | | Rotterdam Study | | | Framingham Study | | |
|---|---|---|---|---|---|---|---|---|---|---|
| | **Taxa** | **Effect $\beta$ (95%CI)** | **P. FE**** | **P. Fisher (direction)#** | **Effect $\beta$ (95%CI)** | **P. adjusted** | **Nnot0** | **Effect $\beta$ (95%CI)** | **P. adjusted** | **Nnot0** |
| Overall | Paraprevotella | 0.26 (0.08, 0.44) | 4.19e-03 | 5.21e-3 (+) | 0.17 (-0.06, 0.40) | 0.48 | 495 | 0.39 (0.11, 0.67) | 0.07 | 311 |
| | Eisenbergiella | -0.13 (-0.23, -0.02) | 0.02 | 0.04 (-) | -0.12 (-0.24, -3.48e-03) | 0.24 | 175 | -0.15 (-0.38, 0.07) | 0.44 | 532 |
| | Hungatella | -0.11 (-0.20, -0.01) | 0.02 | 0.06 (-) | -0.10 (-0.20, 9.34e-04) | 0.26 | 134 | -0.13 (-0.34, 0.08) | 0.50 | 527 |
| | Lachnospiraceae.NK4A136.group | 0.14 (0.02, 0.26) | 0.02 | 0.04 (+) | 0.12 (-0.03, 0.27) | 0.42 | 1114 | 0.17 (-0.03, 0.37) | 0.33 | 1122 |
| | Lachnospiraceae.ND3007.group | 0.15 (0.01, 0.29) | 0.03 | 0.05 (+) | 0.17 (-3.38e-03, 0.35) | 0.27 | 1075 | 0.12 (-0.10, 0.34) | 0.57 | 880 |
| | GCA.900066575 | 0.13 (0.01, 0.24) | 0.03 | 0.04 (+) | 0.09 (-0.05, 0.23) | 0.56 | 434 | 0.20 (1.20e-4, 0.40) | 0.23 | 829 |
| | Roseburia | 0.11 (0.01, 0.21) | 0.04 | 0.04 (+) | 0.08 (-0.04, 0.20) | 0.53 | 1202 | 0.17 (-0.01, 0.35) | 0.27 | 1154 |
| | Lachnospiraceae.UCG.004 | 0.14 (0.01, 0.27) | 0.04 | 0.05 (+) | 0.10 (-0.05, 0.26) | 0.54 | 603 | 0.21 (-0.02, 0.45) | 0.29 | 949 |
| | Flavonifractor | -0.10 (-0.20, -0.00) | 0.05 | 0.02 (-) | -0.04 (-0.17, 0.09) | 0.82 | 309 | -0.22 (-0.38, -0.05) | 0.10 | 1116 |
| | Family.XIII.UCG.001 | 0.11 (-0.00, 0.22) | 0.06 | 0.07 (+) | 0.02 (-0.15, 0.20) | 0.68 | 1122 | 0.18 (0.03, 0.33) | 0.15 | 456 |
| Male | Flavonifractor | -0.19 (-0.32, -0.05) | 5.79e-03 | 0.02 (-) | -0.23 (-0.40, -0.07) | 0.15 | 115 | -0.11 (-0.34, 0.12) | 0.78 | 488 |
| | DTU089 | -0.22 (-0.39, -0.04) | 0.01 | 0.01 (-) | -0.13 (-0.34, 0.09) | 0.71 | 205 | -0.38 (-0.67, -0.09) | 0.19 | 203 |
| | Lachnospiraceae.UCG.010 | 0.20 (0.03, 0.37) | 0.02 | 0.04 (+) | 0.16 (-0.09, 0.41) | 0.67 | 211 | 0.24 (-1.12e-03, 0.48) | 0.42 | 475 |
| | Parasutterella | -0.21 (-0.40, -0.02) | 0.03 | 0.07 (-) | -0.20 (-0.43, 0.04) | 0.55 | 159 | -0.24 (-0.58, 0.11) | 0.63 | 399 |
| | Roseburia | 0.14 (0.01, 0.27) | 0.04 | 0.06 (+) | 0.13 (-0.03, 0.28) | 0.58 | 504 | 0.16 (-0.07, 0.39) | 0.63 | 512 |
| | Subdoligranulum | 0.16 | 0.04 | 0.08 (+) | 0.19 | 0.31 | 505 | 0.04 | 0.96 | 482 |

| | | | | | | | | | | |
|---|---|---|---|---|---|---|---|---|---|---|
| | | (0.01, 0.31) | | | (0.02, 0.36) | | | (-0.29, 0.37) | | |
| | Lachnospiraceae.NK4A136.group | 0.16<br>(-0.00, 0.32) | 0.05 | 0.08 (+) | 0.18<br>(-0.03, 0.39) | 0.53 | 470 | 0.13<br>(-0.12, 0.37) | 0.76 | 496 |
| | Fournierella | 0.13<br>(-0.01, 0.26) | 0.07 | 0.04 (+) | 0.07<br>(-0.09, 0.23) | 0.81 | 109 | 0.30<br>(0.02, 0.57) | 0.36 | 235 |
| | Paraprevotella | 0.21<br>(-0.04, 0.46) | 1.00 | 0.14 (+) | 0.26<br>(-0.08, 0.61) | 0.60 | 229 | 0.15<br>(-0.20, 0.51) | 0.81 | 162 |
| | Family.XIII.UCG.001 | 0.13<br>(-0.03, 0.29) | 0.12 | 0.17 (+) | 0.02<br>(-0.23, 0.27) | 0.99 | 335 | 0.20<br>(-0.01, 0.41) | 0.46 | 206 |
| Female | Lachnospiraceae.UCG.004 | 0.24<br>(0.07, 0.41) | 6.31e-03 | 6.22e-03 (+) | 0.18<br>(-0.02, 0.38) | 0.50 | 352 | 0.40<br>(0.07, 0.73) | 0.21 | 505 |
| | Alistipes | 0.23<br>(0.06, 0.40) | 6.92e-03 | 0.01 (+) | 0.21<br>(-1.27e-04, 0.43) | 0.40 | 678 | 0.26<br>(-0.01, 0.53) | 0.35 | 628 |
| | Parabacteroides | 0.24<br>(0.07, 0.42) | 7.27e-03 | 4.57e-03 (+) | 0.39<br>(0.15, 0.62) | 0.06 | 582 | 0.06<br>(-0.21, 0.33) | 0.87 | 609 |
| | Bilophila | 0.22<br>(0.04, 0.40) | 0.02 | 0.03 (+) | 0.20<br>(-0.04, 0.43) | 0.54 | 197 | 0.25<br>(-0.03, 0.53) | 0.39 | 504 |
| | Dialister | -0.27<br>(-0.52, -0.01) | 0.04 | 0.06 (-) | -0.23<br>(-0.53, 0.07) | 0.60 | 251 | -0.35<br>(-0.83, 0.13) | 0.50 | 289 |
| | GCA.900066575 | 0.11<br>(-0.00, 0.23) | 0.06 | 0.06 (+) | 0.08<br>(-0.05, 0.21) | 0.71 | 260 | 0.22<br>(-0.03, 0.46) | 0.40 | 448 |
| | Paraprevotella | 0.19<br>(-0.01, 0.40) | 0.07 | 0.03 (+) | 0.05<br>(-0.21, 0.30) | 0.93 | 266 | 0.49<br>(0.13, 0.84) | 0.17 | 149 |
| | Eisenbergiella | -0.13<br>(-0.27, 0.01) | 0.07 | 0.08 (-) | -0.10<br>(-0.25, 0.06) | 0.75 | 111 | -0.25<br>(-0.55, 0.05) | 0.44 | 351 |
| | Lactobacillus | 0.14<br>(-0.02, 0.31) | 0.09 | 0.07 (+) | 0.01<br>(-0.22, 0.25) | 0.98 | 155 | 0.27<br>(0.03, 0.50) | 0.26 | 145 |
| | Odoribacter | 0.15<br>(-0.04, 0.34) | 0.12 | 0.18 (+) | 0.13<br>(-0.10, 0.37) | 0.75 | 443 | 0.18<br>(-0.14, 0.50) | 0.63 | 479 |

*P.FE<0.0002 is significance cutoff after multiple testing correction

**P value obtained from meta-analysis with fixed effects (FE) model

### P value obtained from meta-analysis by P value combination following sample size weighted-Fisher approach

**Table S2C. Top 10 trabecular bone score associated taxa; overall and sex stratified.** The joint analysis included participants from two large observational studies: the Rotterdam Study (RS), and the Framingham Heart Study (FHS) with gut microbiome valid samples.

| | | **Joint analysis*** | | | **Rotterdam Study** | | | **Framingham Study** | | |
|---|---|---|---|---|---|---|---|---|---|---|
| | **Taxa** | **Effect $\beta$ (95%CI)** | **P. FE**** | **P. Fisher (direction)#** | **Effect $\beta$ (95%CI)** | **P. adjusted** | **Nnot0** | **Effect $\beta$ (95%CI)** | **P. adjusted** | **Nnot0** |
| Overall | Slackia | 0.12 (0.03, 0.21) | 0.01 | 9.56e-03 (+) | 0.09 (-0.02, 0.19) | 0.40 | 200 | 0.20 (0.03, 0.38) | 0.15 | 212 |
| | Dorea | 0.10 (0.01, 0.19) | 0.02 | 0.04 (+) | 0.12 (0.03, 0.22) | 0.11 | 1204 | -0.01 (-0.23, 0.21) | 0.98 | 898 |
| | Shuttleworthia | 0.11 (5.57e-03, 0.22) | 0.04 | 0.08 (+) | 0.12 (-3.36e-03, 0.24) | 0.28 | 389 | 0.09 (-0.13, 0.32) | 0.72 | 485 |
| | Parabacteroides | 0.14 (-9.84e-04, 0.28) | 0.05 | 0.09 (+) | 0.14 (-0.06, 0.35) | 0.49 | 1009 | 0.14 (-0.06, 0.33) | 0.51 | 933 |
| | Merdibacter | -0.05 (-0.12, 0.02) | 0.13 | 0.17 (-) | -0.04 (-0.12, 0.04) | 0.67 | 174 | -0.09 (-0.22, 0.04) | 0.53 | 181 |
| | Anaerostipes | -0.07 (-0.16, 0.02) | 0.14 | 0.11 (-) | -0.02 (-0.14, 0.09) | 0.89 | 1219 | -0.15 (-0.31, -1.20-e-04) | 0.28 | 982 |
| | Holdemanella | 0.12 (-0.05, 0.28) | 0.15 | 0.26 (+) | 0.09 (-0.15, 0.32) | 0.77 | 479 | 0.15 (-0.08, 0.38) | 0.54 | 206 |
| | Christensenellaceae. R.7.group | -0.12 (-0.28, 0.04) | 0.16 | 0.23 (-) | -0.13 (-0.32, 0.06) | 0.50 | 1125 | -0.08 (-0.37, 0.22) | 0.84 | 808 |
| | Eisenbergiella | -0.06 (-0.16, 0.04) | 0.21 | 0.04 (-) | -0.02 (-0.13, 0.09) | 0.91 | 175 | -0.26 (-0.48, -0.03) | 0.17 | 457 |
| | UBA1819 | 0.07 (-0.04, 0.18) | 0.22 | 0.20 (+) | 0.14 (-2.37e-03, 0.27) | 0.27 | 795 | -0.06 (-0.26, 0.14) | 0.78 | 808 |
| Male | DTU089 | -0.15 (-0.33, 0.02) | 0.08 | 0.06 (-) | -0.08 (-0.29, 0.12) | 0.84 | 205 | -0.33 (-0.65, -0.01) | 0.48 | 169 |
| | Dorea | 0.11 (-0.02, 0.23) | 0.09 | 0.13 (+) | 0.14 (-3.86e-03, 0.27) | 0.42 | 501 | 0.01 (-0.26, 0.28) | 0.99 | 416 |
| | Adlercreutzia | -0.14 (-0.31, 0.03) | 0.10 | 0.05 (-) | -0.07 (-0.27, 0.13) | 0.87 | 109 | -0.34 (-0.66, -0.01) | 0.48 | 359 |
| | Family.XIII.UCG.001 | -0.13 (-0.29, 0.04) | 0.13 | 0.18 (-) | -0.16 (-0.40, 0.08) | 0.64 | 335 | -0.09 (-0.32, 0.13) | 0.87 | 172 |
| | ^Negativibacillus | -0.25 (-0.61, 0.11) | 0.17 | 4.18e-03 (-) | -0.08 (-0.30, 0.13) | 0.86 | 225 | -0.45 (-0.74, -0.16) | 0.14 | 327 |

| | | | | | | | | | | |
|---|---|---|---|---|---|---|---|---|---|---|
| | Shuttleworthia | 0.10<br>(-0.04, 0.23) | 0.17 | 0.22 (+) | 0.07<br>(-0.10, 0.24) | 0.82 | 146 | 0.15<br>(-0.09, 0.39) | 0.78 | 185 |
| | Family.XIII.AD3011.group | -0.10<br>(-0.25, 0.05) | 0.20 | 0.29 (-) | -0.09<br>(-0.26, 0.08) | 0.76 | 469 | -0.13<br>(-0.45, 0.19) | 0.87 | 318 |
| | Butyricimonas | -0.10<br>(-0.25, 0.06) | 0.22 | 0.41 (-) | -0.10<br>(-0.27, 0.06) | 0.69 | 134 | -0.06<br>(-0.42, 0.30) | 0.96 | 270 |
| | Lachnospiraceae.ND3007.group | 0.11<br>(-0.07, 0.28) | 0.23 | 0.21 (+) | 0.20<br>(-0.03, 0.43) | 0.52 | 460 | -0.02<br>(-0.29, 0.25) | 0.98 | 349 |
| | Intestinimonas | -0.10<br>(-0.26, 0.07) | 0.26 | 0.23 (-) | 0.06<br>(-0.22, 0.35) | 0.92 | 305 | -0.18<br>(-0.38, 0.03) | 0.63 | 425 |
| Female | Slackia | 0.15<br>(0.04, 0.25) | 5.00e-03 | 8.58e-03 (+) | 0.13<br>(-3.10e-03, 0.25) | 0.43 | 105 | 0.19<br>(0.01, 0.36) | 0.34 | 93 |
| | Holdemanella | 0.20<br>(4.78e-03, 0.39) | 0.05 | 0.08 (+) | 0.20<br>(-0.05, 0.46) | 0.57 | 251 | 0.19<br>(-0.11, 0.48) | 0.61 | 88 |
| | Parabacteroides | 0.16<br>(-0.01, 0.34) | 0.07 | 0.11 (+) | 0.14<br>(-0.08, 0.37) | 0.73 | 582 | 0.19<br>(-0.09, 0.48) | 0.59 | 510 |
| | Holdemania | 0.11<br>(-0.01, 0.24) | 0.07 | 0.16 (+) | 0.15<br>(3.64e-3, 0.30) | 0.39 | 165 | 0.02<br>(-0.22, 0.26) | 0.96 | 377 |
| | Christensenellaceae.R.7.group | -0.19<br>(-0.40, 0.02) | 0.08 | 0.11 (-) | -0.23<br>(-0.47, 0.01) | 0.44 | 660 | -0.06<br>(-0.47, 0.36) | 0.93 | 445 |
| | Shuttleworthia | 0.11<br>(-0.02, 0.24) | 0.08 | 0.17 (+) | 0.13<br>(-7.20e-03, 0.28) | 0.45 | 243 | 0.01<br>(-0.30, 0.33) | 0.98 | 300 |
| | UC5.1.2E3 | 0.08<br>(-0.02, 0.18) | 0.12 | 0.26 (+) | 0.08<br>(-0.03, 0.19) | 0.60 | 91 | 0.08<br>(-0.23, 0.40) | 0.88 | 221 |
| | Lactobacillus | 0.13<br>(-0.03, 0.30) | 0.12 | 0.17 (+) | 0.09<br>(-0.14, 0.32) | 0.85 | 155 | 0.18<br>(-0.06, 0.41) | 0.55 | 124 |
| | Hungatella | 0.07<br>(-0.02, 0.17) | 0.14 | 0.48 (+) | 0.10<br>(-5.12e-03, 0.20) | 0.45 | 85 | -0.13<br>(-0.42, 0.17) | 0.76 | 272 |
| | Dorea | 0.08<br>(-0.03, 0.20) | 0.15 | 0.25 (+) | 0.10<br>(-0.02, 0.22) | 0.58 | 703 | -0.01<br>(-0.34, 0.31) | 0.98 | 482 |

*P<0.0002 is significance cutoff after multiple testing correction
**P value obtained from meta-analysis with fixed effects (FE) model
### P value obtained from meta-analysis by P value combination following sample size weighted-Fisher approach
^ The taxon in red color indicates that a random effects model was used rather than FE model

**TableS3A. Genera associated with appendicular lean mass at P.adjusted <0.1 within each study cohort; overall and sex-stratified.**

| | Rotterdam Study | | | | Framingham Study | | | |
|---|---|---|---|---|---|---|---|---|
| | **Taxa** | **Effect $\beta$ (95%CI)** | **P.adjusted** | **Nnot0** | **Taxa** | **Effect $\beta$ (95%CI)** | **P.adjusted** | **Nnot0** |
| Overall | Oscillibacter | -0.62 (-0.94, -0.30) | 5.80e-03 | 690 | Howardella | 0.51 (0.26, 0.76) | 5.60e-03 | 161 |
| | Alistipes | -0.60 (-0.94, -0.26) | 0.02 | 1145 | Romboutsia | 0.92 (0.39, 1.45) | 0.02 | 664 |
| | Eisenbergiella | -0.37 (-0.59, -0.15) | 0.02 | 175 | Subdoligranulum | 0.84 (0.30, 1.38) | 0.05 | 985 |
| | Agathobacter | 0.40 (0.16, 0.64) | 0.03 | 1192 | Lachnospira | 0.55 (0.19, 0.91) | 0.05 | 1043 |
| | Prevotella_9 | 1.09 (0.43, 1.75) | 0.03 | 447 | Faecalibacterium | 0.56 (0.20, 0.92) | 0.05 | 1063 |
| | Family.XIII.AD3011.group | -0.39 (-0.63, -0.15) | 0.03 | 1122 | Collinsella | 0.70 (0.24, 1.17) | 0.05 | 858 |
| | UCG.007 | -0.27 (-0.43, -0.10) | 0.03 | 193 | Sellimonas | -0.64 (-1.05, -0.22) | 0.05 | 535 |
| | Anaerotruncus | -0.35 (-0.58, -0.13) | 0.04 | 241 | Eggerthella | -0.63 (-1.05, -0.21) | 0.05 | 565 |
| | Streptococcus | 0.65 (0.22, 1.08) | 0.05 | 1009 | Anaerotruncus | -0.61 (-1.02, -0.20) | 0.06 | 743 |
| | Hungatella | -0.27 (-0.47, -0.08) | 0.07 | 134 | Defluviitaleaceae_UCG.011 | 0.48 (0.14, 0.82) | 0.08 | 349 |
| | DTU089 | -0.38 (-0.66, -0.10) | 0.09 | 509 | Faecalitalea | -0.64 (-1.10, -0.18) | 0.09 | 427 |
| | Bacteroides | -0.35 (-0.62, -0.09) | 0.09 | 1234 | Lachnospiraceae_UCG.008 | 0.51 (0.14, 0.89) | 0.09 | 257 |
| | Erysipelatoclostridium | -0.45 (-0.79, -0.12) | 0.09 | 467 | Coprococcus_1 | 0.51 (0.13, 0.90) | 0.09 | 649 |
| | | | | | CAG.56 | 0.71 (0.18, 1.24) | 0.09 | 574 |
| | | | | | Roseburia | 0.51 (0.13, 0.89) | 0.09 | 1066 |
| | | | | | Coprococcus_3 | 0.74 (0.18, 1.30) | 0.10 | 851 |

| | | | | | | | | |
|---|---|---|---|---|---|---|---|---|
| Male | Oscillibacter | -0.52<br>(-0.79, -0.24) | 0.03 | 267 | No genera were identified at P.adjusted<0.1 | | | |
| Female | Hungatella | -0.26<br>(-0.38, -0.13) | 0.01 | 85 | Sellimonas | -0.56<br>(-0.86, -0.27) | 0.04 | 336 |
| | Anaerotruncus | -0.28<br>(-0.42, -0.14) | 0.01 | 156 | Lachnospiraceae_UCG.010 | 0.43<br>(0.19, 0.66) | 0.06 | 579 |
| | Escherichia.Shigella | -0.66<br>(-1.07, -0.24) | 0.08 | 501 | | | | |
| | Family.XIII.AD3011.group | -0.29<br>(-0.47, -0.10) | 0.08 | 653 | | | | |
| | DTU089 | -0.33<br>(-0.54, -0.12) | 0.08 | 304 | | | | |

**TableS3B. Genera associated with femoral neck bone mineral density at P.adjusted<0.1 within each study cohort; overall and sex-stratified.**

| | Rotterdam Study | | | | Framingham Study | | | |
|---|---|---|---|---|---|---|---|---|
| | **Taxa** | **Effect $\beta$ (95%CI)** | **P.adjusted** | **Nnot0** | **Taxa** | **Effect $\beta$ (95%CI)** | **P.adjusted** | **Nnot0** |
| Overall | Eubacterium.eligens.group | 0.29 (0.09, 0.50) | 0.07 | 973 | DTU089 | -0.45 (-0.68, -0.22) | 3.70e-03 | 537 |
| | | | | | Faecalitalea | -0.35 (-0.57, -0.13) | 0.03 | 467 |
| | | | | | Paraprevotella | 0.39 (0.11, 0.67) | 0.07 | 311 |
| | | | | | Ruminococcaceae_UCG.007 | 0.18 (0.05, 0.32) | 0.08 | 192 |
| Male | No genera were identified at P.adjusted<0.1 | | | | Ruminococcaceae_UCG.007 | 0.28 (0.10, 0.45) | 0.09 | 67 |
| Female | Parabacteroides | 0.39 (0.15, 0.62) | 0.06 | 582 | Lachnospiraceae_UCG.008 | 0.33 (0.13, 0.53) | 0.08 | 148 |
| | Eubacterium.eligens.group | 0.42 (0.15, 0.68) | 0.07 | 578 | | | | |

**TableS3C. Genera associated with trabecular bone score at P.adjusted<0.1 within each study cohort; overall and sex-stratified.**

| | Rotterdam Study | | | | Framingham Study | | | |
|---|---|---|---|---|---|---|---|---|
| | **Taxa** | **Effect β (95%CI)** | **P.adjusted** | **Nnot0** | **Taxa** | **Effect β (95%CI)** | **P.adjusted** | **Nnot0** |
| Overall | UCG.007 | -0.13 (-0.21, -0.05) | 0.02 | 193 | Sellimonas | -0.31 (-0.51, -0.11) | 0.03 | 497 |
| | | | | | Blautia | -0.16 (-0.27, -0.05) | 0.06 | 1008 |
| | | | | | DTU089 | -0.33 (-0.56, -0.10) | 0.06 | 458 |
| Male | UCG.007 | -0.17 (-0.28, -0.07) | 0.07 | 69 | No genera were identified at P.adjusted<0.1 | | | |
| Female | Butyrivibrio | -0.34 (-0.55, -0.12) | 0.09 | 121 | No genera were identified at P.adjusted<0.1 | | | |

**Table S4A. Top 10 Appendicular lean mass associated pathway; overall and sex-stratified**. The joint analysis included participants from two large observational studies: the Rotterdam Study (RS), and the Framingham Heart Study (FHS) with gut microbiome valid samples.

| | | | **Joint analysis*** | | | **Rotterdam Study** | | | **Framingham Study** | | |
|---|---|---|---|---|---|---|---|---|---|---|---|
| | **Pathway** | **Pathway description** | **Effect β (95%CI)** | **P. FE**** | **P. Fisher (direction) #** | **Effect β (95%CI)** | **P. adjusted** | **Nnot0** | **Effect β (95%CI)** | **P. adjusted** | **Nnot0** |
| Overall | PWY.5005 | Biotin biosynthesis II | 0.44 (0.17, 0.70) | 1.19e-03 | 2.31e-03 (+) | 0.39 (0.01, 0.76) | 0.20 | 1131 | 0.49 (0.11, 0.86) | 0.16 | 1065 |
| | CENTFERM.PWY | Pyruvate fermentation to butanoate | -0.24 (-0.40, -0.09) | 1.73e-03 | 3.44e-03 (-) | -0.26 (-0.47, -0.04) | 0.12 | 1225 | -0.23 (-0.45, -0.01) | 0.27 | 1102 |
| | PYRIDOXSYN.PWY | Pyridoxal 5'-phosphate biosynthesis I | -0.18 (-0.30, -0.06) | 2.66e-03 | 2.21e-03 (-) | -0.32 (-0.55, -0.08) | 0.07 | 1242 | -0.14 (-0.27, 2.20e-03) | 0.30 | 1105 |
| | PWY.5532 | Adenosine nucleotides degradation IV | -0.31 (-0.51, -0.11) | 2.76e-03 | 2.92e-03 (-) | -0.24 (-0.47, -0.02) | 0.18 | 233 | -0.55 (-0.99, -0.11) | 0.19 | 659 |
| | PWY.5913 | TCA cycle VI (obligate autotrophs) | -0.12 (-0.20, -0.04) | 4.03e-03 | 4.24e-03 (-) | -0.05 (-0.17, 0.08) | 0.69 | 1246 | -0.16 (-0.27, -0.06) | 0.07 | 1105 |
| | P163.PWY | L-lysine fermentation to acetate and butanoate | -0.19 (-0.32, -0.06) | 4.07e-03 | 7.73e-03 (-) | -0.21 (-0.41, -3.15e-03) | 0.20 | 1198 | -0.18 (-0.35, -0.01) | 0.27 | 1101 |
| | PWY.5028 | L-histidine degradation II | -0.34 (-0.58, -0.10) | 4.86e-03 | 5.20e-03 (-) | -0.27 (-0.54, 8.01e-03) | 0.23 | 168 | -0.58 (-1.07, -0.10) | 0.21 | 470 |
| | PWY.6641 | Superpathway of sulfolactate degradation | -0.28 (-0.48, -0.09) | 4.90e-03 | 0.02 (-) | -0.33 (-0.54, -0.12) | 0.03 | 167 | 0.01 (-0.51, 0.53) | 0.98 | 475 |
| | PWY.6628 | Superpathway of L-phenylalanine biosynthesis | 0.11 (0.03, 0.19) | 6.95e-03 | 0.01 (+) | 0.10 (-0.02, 0.22) | 0.30 | 1244 | 0.12 (9.32e-03, 0.24) | 0.26 | 1105 |

| | | | | | | | | | | | |
|---|---|---|---|---|---|---|---|---|---|---|---|
| | PWY.7013 | L-1,2-propanediol degradation | -0.22 (-0.38, -0.06) | 7.32e-03 | 0.01 (-) | -0.21 (-0.44, 0.03) | 0.30 | 1169 | -0.23 (-0.45, -0.01) | 0.27 | 1095 |
| Male | PWY.6641 | Superpathway of sulfolactate degradation | -0.24 (-0.40, -0.09) | 2.23e-03 | 0.01 (-) | -0.27 (-0.44, -0.10) | 0.05 | 66 | -0.08 (-0.48, 0.31) | 0.91 | 200 |
| | PWY.4984 | Urea cycle | -0.17 (-0.28, -0.06) | 3.44e-03 | 2.89e-03 (-) | -0.10 (-0.25, 0.05) | 0.63 | 521 | -0.27 (-0.44, -0.09) | 0.14 | 462 |
| | PWY.7328 | Superpathway of UDP-glucose-derived O-antigen building blocks biosynthesis | -0.12 (-0.22, -0.02) | 0.02 | 0.03 (-) | -0.12 (-0.29, 0.06) | 0.60 | 516 | -0.13 (-0.25, -2.52e-03) | 0.38 | 462 |
| | CENTFERM.PWY | Pyruvate fermentation to butanoate | -0.16 (-0.31, -0.02) | 0.03 | 0.04 (-) | -0.11 (-0.32, 0.10) | 0.71 | 505 | -0.21 (-0.40, -0.01) | 0.34 | 460 |
| | PRPP.PWY | Superpathway of histidine, purine, and pyrimidine biosynthesis | -0.08 (-0.15, -9.17e-03) | 0.03 | 0.03 (-) | -0.15 (-0.32, 0.02) | 0.43 | 513 | -0.06 (-0.14, 0.01) | 0.51 | 462 |
| | PWY.5532 | Adenosine nucleotides degradation IV | -0.14 (-0.27, -9.65e-03) | 0.04 | 0.03 (-) | -0.11 (-0.25, 0.03) | 0.51 | 89 | -0.32 (-0.66, 0.02) | 0.44 | 281 |
| | PWY.6353 | Purine nucleotides degradation II (aerobic) | -0.05 (-0.09, -1.45e-03) | 0.04 | 0.06 (-) | -0.03 (-0.09, 0.03) | 0.69 | 521 | -0.07 (-0.14, 3.52e-03) | 0.44 | 461 |
| | PWY490.3 | Nitrate reduction VI (assimilatory) | 0.07 (1.59e-03, 0.14) | 0.05 | 0.06 (+) | 0.05 (-0.04, 0.14) | 0.67 | 521 | 0.11 (-9.64e-03, 0.22) | 0.45 | 462 |

| | | | | | | | | | | | |
|---|---|---|---|---|---|---|---|---|---|---|---|
| | PWY.5913 | TCA cycle VI (obligate autotrophs) | -0.07 (-0.14, -8.89e-04) | 0.05 | 0.05 (-) | -9.00e-03 (-0.12, 0.10) | 0.97 | 518 | -0.11 (-0.20, -0.02) | 0.27 | 462 |
| | P163.PWY | L-lysine fermentation to acetate and butanoate | -0.11 (-0.22, 2.72e-03) | 0.06 | 0.08 (-) | -0.16 (-0.36, 0.05) | 0.50 | 493 | -0.09 (-0.23, 0.05) | 0.63 | 460 |
| Female | PWY.5005 | Biotin biosynthesis II | 0.44 (0.24, 0.64) | 1.55e-05 | 3.95e-05 (+) | 0.42 (0.13, 0.71) | 0.08 | 664 | 0.46 (0.18, 0.73) | 0.11 | 617 |
| | PWY.6628 | Superpathway of L-phenylalanine biosynthesis | 0.11 (0.04, 0.17) | 9.65e-04 | 2.03e-03 (+) | 0.11 (0.01, 0.20) | 0.22 | 725 | 0.11 (0.02, 0.19) | 0.28 | 643 |
| | PWY.5028 | L-histidine degradation II | -0.3 (-0.49, -0.11) | 1.51e-03 | 4.76e-03 (-) | -0.33 (-0.56, -0.10) | 0.09 | 107 | -0.25 (-0.57, 0.06) | 0.58 | 296 |
| | POLYAMINSYN3.PWY | Superpathway of polyamine biosynthesis II | 0.15 (0.05, 0.25) | 2.07e-03 | 4.08e-03 (+) | 0.08 (-0.10, 0.27) | 0.73 | 714 | 0.18 (0.06, 0.29) | 0.15 | 643 |
| | P125.PWY | Superpathway of (R,R)-butanediol biosynthesis | -0.2 (-0.35, -0.06) | 6.43e-03 | 2.84e-03 (-) | -0.13 (-0.30, 0.03) | 0.45 | 722 | -0.46 (-0.77, -0.14) | 0.21 | 545 |
| | PWY.7013 | L-1,2-propanediol degradation | -0.13 (-0.24, -0.03) | 0.01 | 0.02 (-) | -0.14 (-0.29, 0.01) | 0.32 | 690 | -0.13 (-0.28, 0.02) | 0.54 | 640 |
| | AST.PWY | L-arginine degradation II (AST pathway) | -0.39 (-0.69, -0.08) | 0.01 | 5.30e-03 (-) | -0.67 (-1.10, -0.25) | 0.05 | 517 | -0.09 (-0.53, 0.35) | 0.92 | 370 |
| | PWY.6470 | Peptidoglycan biosynthesis V (β-lactam resistance) | 0.26 (0.05, 0.47) | 0.02 | 0.02 (+) | 0.36 (0.05, 0.68) | 0.20 | 632 | 0.17 (-0.11, 0.45) | 0.69 | 581 |
| | P341.PWY | Glycolysis V (Pyrococcus) | -0.26 (-0.47, -0.05) | 0.02 | 0.02 (-) | -0.18 (-0.45, 0.09) | 0.56 | 242 | -0.39 (-0.74, -0.04) | 0.34 | 497 |

| | | | | | | | | | | | |
|---|---|---|---|---|---|---|---|---|---|---|---|
| | PWY.7392 | Taxadiene biosynthesis (engineered) | 0.14 (0.02, 0.25) | 0.02 | 0.02 (+) | 0.09 (-0.06, 0.24) | 0.60 | 723 | 0.19 (0.02, 0.36) | 0.34 | 640 |

*P<0.00019 is significance cutoff after multiple testing correction
**P value obtained from meta-analysis with fixed effects (FE) model
### P value obtained from meta-analysis by P value combination following sample size weighted-Fisher approach

**Table S4B. Top 10 Femoral neck bone mineral density associated pathway; overall and sex stratified**. The joint analysis included participants from two large observational studies: the Rotterdam Study (RS), and the Framingham Heart Study (FHS) with gut microbiome valid samples.

| | | | Joint analysis | | | Rotterdam Study | | | Framingham Study | | |
|---|---|---|---|---|---|---|---|---|---|---|---|
| | **Pathway** | **Pathway description** | **Effect β (95%CI)** | **P. FE**** | **P. Fisher (direction) #** | **Effect β (95%CI)** | **P. adjusted** | **Nnot0** | **Effect β (95%CI)** | **P. adjusted** | **Nnot0** |
| Overall | P163.PWY | L-lysine fermentation to acetate and butanoate | -0.07 (-0.14, -0.01) | 0.03 | 0.03 (-) | -0.02 (-0.13, 0.09) | 0.89 | 1198 | -0.10 (-0.18, -0.02) | 0.13 | 1192 |
| | PWY.7371 | 1,4-dihydroxy-6-naphthoate biosynthesis II | 0.12 (0.01, 0.23) | 0.04 | 0.05 (+) | 0.17 (0.02, 0.32) | 0.16 | 1188 | 0.06 (-0.10, 0.22) | 0.78 | 1176 |
| | CENTFERM.PWY | Pyruvate fermentation to butanoate | -0.07 (-0.15, 2.18E-03) | 0.06 | 0.06 (-) | -0.12 (-0.23, -9.19e-03) | 0.16 | 1225 | -0.03 (-0.14, 0.07) | 0.82 | 1194 |
| | PWY0.781 | Aspartate superpathway | -0.02 (-0.04, 1.37E-03) | 0.07 | 0.12 (-) | -0.02 (-0.05, 7.31e-05) | 0.21 | 1249 | -0.01 (-0.05, 0.04) | 0.95 | 1197 |
| | PWY.6478 | GDP-D-glycero-α-D-manno-heptose biosynthesis | 0.03 (-0.01, 0.07) | 0.09 | 0.12 (+) | 0.02 (-0.04, 0.07) | 0.79 | 1249 | 0.05 (-4.88e-03, 0.10) | 0.36 | 1197 |
| | PWY.5177 | Glutaryl-CoA degradation | -0.06 (-0.13, 0.02) | 0.14 | 0.18 (-) | -0.09 (-0.20, 0.03) | 0.40 | 1221 | -0.03 (-0.14, 0.07) | 0.81 | 1194 |
| | PWY.7013 | L-1,2-propanediol degradation | -0.05 (-0.13, 0.03) | 0.19 | 0.20 (-) | 7.31e-03 (-0.12, 0.13) | 0.96 | 1169 | -0.10 (-0.20, 8.88e-03) | 0.35 | 1186 |
| | PWY.7254 | TCA cycle VII (acetate-producers) | -0.05 (-0.13, 0.03) | 0.20 | 0.24 (-) | -0.01 (-0.13, 0.11) | 0.94 | 1229 | -0.08 (-0.19, 0.02) | 0.44 | 1195 |
| | POLYAMINSYN3.PWY | Superpathway of polyamine biosynthesis II | 0.04 (-0.02, 0.10) | 0.22 | 0.34 (+) | 0.03 (-0.09, 0.15) | 0.83 | 1225 | 0.04 (-0.03, 0.11) | 0.61 | 1196 |
| | LEU.DEG2.PWY | L-leucine degradation I | 0.11 (-0.08, 0.30) | 0.24 | 0.35 (+) | 0.14 (-0.09, 0.38) | 0.51 | 530 | 0.06 (-0.26, 0.37) | 0.91 | 537 |

| | | | | | | | | | | | |
|---|---|---|---|---|---|---|---|---|---|---|---|
| Male | PWY.7198 | Pyrimidine deoxyribonucleotides de novo biosynthesis IV | 0.08 (0.02 , 0.14) | 0.01 | 0.02 (+) | 0.09 (0.02, 0.16) | 0.14 | 518 | 0.04 (-0.09, 0.16) | 0.9 | 528 |
| | P163.PWY | L-lysine fermentation to acetate and butanoate | -0.10 (-0.20, -0.01) | 0.03 | 0.05 (-) | -0.15 (-0.32, 0.02) | 0.43 | 493 | -0.08 (-0.20, 0.03) | 0.65 | 527 |
| | PWY.7254 | TCA cycle VII (acetate-producers) | -0.10 (-0.20, 4.28e-03) | 0.06 | 0.09 (-) | -0.06 (-0.22, 0.10) | 0.84 | 511 | -0.13 (-0.27, 8.20e-03) | 0.43 | 530 |
| | ARGORNPROST.PWY | Arginine, ornithine and proline interconversion | 0.05 (-3.17e-03, 0.11) | 0.07 | 0.05 (+) | 0.09 (0.02, 0.17) | 0.21 | 521 | 0.01 (-0.07, 0.09) | 0.97 | 530 |
| | PWY.5677 | Succinate fermentation to butanoate | -0.06 (-0.13, 4.61e-03) | 0.07 | 0.12 (-) | -0.03 (-0.19, 0.13) | 0.95 | 501 | -0.07 (-0.15, 3.44e-03) | 0.43 | 529 |
| | PWY.5022 | 4-aminobutanoate degradation V | -0.09 (-0.18, 0.01) | 0.07 | 0.12 (-) | -0.07 (-0.21, 0.07) | 0.74 | 515 | -0.10 (-0.22, 0.03) | 0.60 | 527 |
| | PWY.7328 | Superpathway of UDP-glucose-derived O-antigen building blocks biosynthesis | -0.08 (-0.16, 0.01) | 0.08 | 0.10 (-) | -8.56e-03 (-0.16, 0.14) | 0.99 | 516 | -0.11 (-0.21, -4.12e-03) | 0.36 | 530 |
| | PWY.7234 | Inosine-5'-phosphate biosynthesis III | -0.05 (-0.12, 0.01) | 0.11 | 0.10 (-) | -0.10 (-0.21, 7.45e-03) | 0.39 | 519 | -0.03 (-0.11, 0.05) | 0.88 | 530 |
| | PWY.7392 | Taxadiene biosynthesis (engineered) | -0.09 (-0.18, 0.02) | 0.13 | 0.18 (-) | -0.11 (-0.25, 0.03) | 0.51 | 515 | -0.04 (-0.19, 0.11) | 0.90 | 529 |
| | AST.PWY | TCA cycle VII (acetate-producers) | -0.20 (-0.46, 0.07) | 0.15 | 0.14 (-) | -0.39 (-0.85, 0.08) | 0.47 | 347 | -0.10 (-0.43, 0.22) | 0.88 | 322 |

| | | | | | | | | | | | |
|---|---|---|---|---|---|---|---|---|---|---|---|
| Female | CENTFERM.PWY | Pyruvate fermentation to butanoate | -0.14 (-0.25, -0.04) | 9.23e-03 | 0.01 (-) | -0.22 (-0.40, -0.04) | 0.21 | 505 | -0.10 (-0.23, 0.03) | 0.66 | 666 |
| | PWY.5177 | Glutaryl-CoA degradation | -0.13 (-0.24, -0.02) | 0.02 | 0.02 (-) | -0.21 (-0.39, -0.02) | 0.26 | 503 | -0.09 (-0.23, 0.04) | 0.68 | 666 |
| | P163.PWY | L-lysine fermentation to acetate and butanoate | -0.11 (-0.20, -0.02) | 0.02 | 0.03 (-) | -0.15 (-0.32, 0.02) | 0.43 | 493 | -0.09 (-0.20, 0.02) | 0.61 | 665 |
| | PWY.5028 | L-histidine degradation II | -0.16 (-0.31, -4.12e-03) | 0.04 | 0.03 (-) | -0.09 (-0.27, 0.09) | 0.74 | 61 | -0.31 (-0.59, -0.03) | 0.38 | 308 |
| | PWY.6471 | Peptidoglycan biosynthesis IV (Enterococcus faecium) | 0.05 (1.08e-03, 0.09) | 0.05 | 0.06 (+) | 0.07 (5.00e-03, 0.13) | 0.29 | 521 | 0.02 (-0.04, 0.09) | 0.87 | 667 |
| | PWY.5920 | Superpathway of heme biosynthesis from glycine | -0.25 (-0.51, 0.02) | 0.07 | 0.06 (-) | -0.45 (-0.82, -0.08) | 0.21 | 393 | -0.03 (-0.41, 0.36) | 0.98 | 579 |
| | PWY490.3 | Nitrate reduction VI (assimilatory) | 0.05 (-0.01, 0.11) | 0.12 | 0.20 (+) | 0.05 (-0.03, 0.12) | 0.64 | 521 | 0.05 (-0.06, 0.15) | 0.82 | 667 |
| | GLUCARGALACTSUPER.PWY | Superpathway of D-glucarate and D-galactarate degradation | -0.16 (-0.36, 0.04) | 0.12 | 0.15 (-) | -0.08 (-0.39, 0.23) | 0.91 | 341 | -0.21 (-0.48, 0.05) | 0.63 | 626 |
| | PWY.6891 | Thiazole biosynthesis II (Bacillus) | 0.03 (-0.01, 0.08) | 0.13 | 0.23 (+) | 0.03 (-0.01, 0.08) | 0.58 | 521 | 0.03 (-0.07, 0.12) | 0.91 | 667 |
| | PWY.5507 | Adenosylcobalamin biosynthesis I (early cobalt insertion) | -0.14 (-0.31, 0.04) | 0.14 | 0.14 (-) | -0.03 (-0.29, 0.23) | 0.97 | 163 | -0.24 (-0.48, 0.01) | 0.52 | 161 |

*P<0.00019 is significance cutoff after multiple testing correction
**P value obtained from meta-analysis with fixed effects (FE) model
### P value obtained from meta-analysis by P value combination following sample size weighted-Fisher approach

**Table S4C. Top 10 Trabecular bone score associated pathway; overall and sex stratified.** The joint analysis included participants from two large observational studies: the Rotterdam Study (RS), and the Framingham Heart Study (FHS) with gut microbiome valid samples.

| | | | Joint analysis | | | Rotterdam Study | | | Framingham Study | | |
|---|---|---|---|---|---|---|---|---|---|---|---|
| | **Pathway** | **Pathway description** | **Effect *β* (95%CI)** | **P. FE**** | **P. Fisher (direction)#** | **Effect *β* (95%CI)** | **P. adjusted** | **Nnot0** | **Effect *β* (95%CI)** | **P. adjusted** | **Nnot0** |
| Overall | POLYAMSYN.PWY | Superpathway of polyamine biosynthesis I | 0.06 (0.01, 0.11) | 0.03 | 0.02 (+) | 0.01 (-0.07, 0.09) | 0.89 | 1244 | 0.10 (0.03, 0.17) | 0.09 | 1011 |
| | POLYAMINSYN3.PWY | Superpathway of polyamine biosynthesis II | 0.07 (0.01, 0.13) | 0.03 | 0.06 (+) | 0.04 (-0.08, 0.15) | 0.77 | 1225 | 0.08 (7.44e-03, 0.15) | 0.21 | 1010 |
| | PWY0.1261 | Anhydromuropeptides recycling | 0.06 (-0.01, 0.14) | 0.09 | 0.08 (+) | 4.91e-03 (-0.10, 0.11) | 0.98 | 1229 | 0.11 (0.02, 0.21) | 0.19 | 1010 |
| | PWY.6628 | Superpathway of L-phenylalanine biosynthesis | 0.03 (-0.01, 0.07) | 0.10 | 0.14 (+) | 0.02 (-0.04, 0.07) | 0.79 | 1244 | 0.05 (-6.88e-03, 0.10) | 0.38 | 1011 |
| | PWY.5265 | Peptidoglycan biosynthesis II (staphylococci) | 0.15 (-0.04, 0.34) | 0.12 | 0.16 (+) | 0.08 (-0.18, 0.34) | 0.78 | 638 | 0.23 (-0.04, 0.50) | 0.40 | 410 |
| | GLYCOLYSIS.E.D | Superpathway of glycolysis and Entner-Doudoroff | 0.04 (-0.01, 0.09) | 0.12 | 0.23 (+) | -1.95e-03 (-0.14, 0.13) | 0.99 | 1210 | 0.05 (-6.92e-03, 0.10) | 0.39 | 1011 |
| | TYRFUMCAT.PWY | L-tyrosine degradation I | -0.08 (-0.19, 0.02) | 0.12 | 0.29 (-) | -0.10 (-0.21, 0.01) | 0.30 | 154 | 0.01 (-0.25, 0.27) | 0.99 | 300 |

| | | | | | | | | | | | |
|---|---|---|---|---|---|---|---|---|---|---|---|
| | HEMESYN2.PWY | Heme biosynthesis II (anaerobic) | 0.05 (-0.01, 0.11) | 0.13 | 0.07 (+) | -5.02e-03 (-0.09, 0.08) | 0.97 | 1244 | 0.11 (0.02, 0.21) | 0.16 | 1010 |
| | P341.PWY | Glycolysis V (Pyrococcus) | -0.11 (-0.24, 0.03) | 0.14 | 0.15 (-) | -0.07 (-0.24, 0.11) | 0.73 | 423 | -0.18 (-0.41, 0.06) | 0.48 | 724 |
| | PWY.5705 | Allantoin degradation to glyoxylate III | 0.04 (-0.01, 0.10) | 0.14 | 0.14 (+) | 0.07 (-0.6.36e-03, 0.15) | 0.26 | 1231 | 0.01 (-0.07, 0.09) | 0.95 | 1011 |
| Male | PWY.7090 | UDP-2,3-diacetamido-2,3-dideoxy-α-D-mannuronate biosynthesis | -0.20 (-0.36, -0.04) | 0.01 | 0.02 (-) | -0.08 (-0.36, 0.20) | 0.89 | 387 | -0.26 (-0.46, -0.07) | 0.32 | 431 |
| | PWY.5705 | Allantoin degradation to glyoxylate III | 0.09 (1.56e-03, 0.18) | 0.05 | 0.07 (+) | 0.11 (-0.02, 0.23) | 0.45 | 510 | 0.07 (-0.05, 0.19) | 0.71 | 450 |
| | NAD.BIOSYNTHESIS.II | NAD salvage pathway II | 0.07 (-0.01, 0.15) | 0.07 | 0.12 (+) | 0.08 (-0.02, 0.18) | 0.47 | 517 | 0.06 (-0.08, 0.19) | 0.83 | 450 |
| | PWY.7013 | L-1,2-propanediol degradation | 0.09 (-0.03, 0.21) | 0.14 | 0.10 (+) | 0.19 (3.92e-03, 0.38) | 0.32 | 479 | 0.02 (-0.13, 0.17) | 0.95 | 443 |
| | POLYAMINSYN3.PWY | Superpathway of polyamine biosynthesis II | 0.06 (-0.02, 0.13) | 0.15 | 0.27 (+) | 0.01 (-0.15, 0.18) | 0.99 | 511 | 0.07 (-0.02, 0.15) | 0.62 | 450 |
| | POLYAMSYN.PWY | Superpathway of polyamine biosynthesis I | 0.05 (-0.02, 0.13) | 0.16 | 0.20 (+) | 3.40e-03 (-0.11, 0.12) | 0.99 | 518 | 0.09 (-9.08e-03, 0.18) | 0.52 | 450 |
| | PWY.7234 | Inosine-5'-phosphate biosynthesis III | 0.04 (-0.02, 0.11) | 0.18 | 0.18 (+) | -0.01 (-0.12, 0.09) | 0.96 | 519 | 0.08 (-2.80e-04, 0.17) | 0.44 | 450 |

| | | | | | | | | | | |
|---|---|---|---|---|---|---|---|---|---|---|
| | P261.PWY | Coenzyme M biosynthesis I | -0.08 (-0.19, 0.04) | 0.19 | 0.21 (-) | -0.06 (-0.18, 0.06) | 0.72 | 64 | -0.20 (-0.57, 0.16) | 0.73 | 151 |
| | PWY.7377 | Cob(II)yrinate a,c-diamide biosynthesis I (early cobalt insertion) | 0.12 (-0.06, 0.29) | 0.19 | 0.25 (+) | 0.09 (-0.12, 0.30) | 0.81 | 502 | 0.18 (-0.13, 0.48) | 0.72 | 432 |
| | LEU.DEG 2.PWY | L-leucine degradation I | 0.14 (-0.07, 0.35) | 0.19 | 0.20 (+) | 0.08 (-0.18, 0.33) | 0.89 | 211 | 0.27 (-0.09, 0.63) | 0.63 | 199 |
| Female | P125.PWY | Superpathway of (R,R)-butanediol biosynthesis | -0.13 (-0.26, -0.01) | 0.03 | 0.05 (-) | -0.12 (-0.26, 0.01) | 0.40 | 722 | -0.20 (-0.49, 0.10) | 0.70 | 471 |
| | P341.PWY | Glycolysis V (Pyrococcus) | -0.19 (-0.37, -5.15e-03) | 0.04 | 0.04 (-) | -0.12 (-0.34, 0.10) | 0.64 | 242 | -0.33 (-0.65, -2.60e-03) | 0.48 | 434 |
| | PWY.5532 | Adenosine nucleotides degradation IV | -0.13 (-0.26, 7.16e-03) | 0.06 | 0.16 (-) | -0.14 (-0.29, 7.83e-03) | 0.35 | 144 | -0.07 (-0.36, 0.23) | 0.94 | 331 |
| | PWY0.126 1 | Anhydromuro peptides recycling | 0.09 (-5.77e-03, 0.19) | 0.07 | 0.10 (+) | 0.07 (-0.06, 0.21) | 0.63 | 718 | 0.11 (-0.03, 0.25) | 0.63 | 561 |
| | PWY.7003 | Glycerol degradation to butanol | -0.15 (-0.33, 0.02) | 0.08 | 0.08 (-) | -0.07 (-0.29, 0.15) | 0.81 | 675 | -0.30 (-0.58, -8.84e-03) | 0.45 | 500 |
| | P162.PWY | L-glutamate degradation V (via hydroxyglutarate) | 0.13 (-0.02, 0.29) | 0.09 | 0.13 (+) | 0.16 (-0.03, 0.36) | 0.44 | 621 | 0.08 (-0.17, 0.34) | 0.91 | 502 |
| | PWY.5507 | Adenosylcobalamin biosynthesis I (early cobalt insertion) | -0.14 (-0.30, 0.03) | 0.10 | 0.12 (-) | -0.03 (-0.26, 0.20) | 0.93 | 263 | -0.25 (-0.50, -0.01) | 0.43 | 129 |

| | | | | | | | | | | | |
|---|---|---|---|---|---|---|---|---|---|---|---|
| | GLYCOLYSIS.E.D | Superpathway of glycolysis and Entner-Doudoroff | 0.06 (-0.01, 0.13) | 0.11 | 0.11 (+) | 0.13 (-0.04, 0.30) | 0.5 | 707 | 0.04 (-0.03, 0.12) | 0.76 | 561 |
| | PWY0.781 | Aspartate superpathway | -0.02 (-0.05, 5.16e-03) | 0.11 | 0.17 (-) | -0.03 (-0.06, 2.25e-03) | 0.36 | 728 | 6.00e-03 (-0.06, 0.07) | 0.98 | 561 |
| | POLYAMINSYN3.PWY | Superpathway of polyamine biosynthesis II | 0.07 (-0.02, 0.16) | 0.13 | 0.21 (+) | 0.04 (-0.11, 0.19) | 0.85 | 714 | 0.08 (-0.03, 0.19) | 0.63 | 560 |

*P<0.00019 is significance cutoff after multiple testing correction

**P value obtained from meta-analysis with fixed effects (FE) model

### P value obtained from meta-analysis by P value combination following sample size weighted-Fisher approach

**TableS5A. Pathways associated with appendicular lean mass at P.adjusted<0.1 within each study cohort; overall and sex-stratified**.

| | Rotterdam Study | | | | | Framingham Study | | | | |
|---|---|---|---|---|---|---|---|---|---|---|
| | **Pathway** | **Pathway description** | **Effect $\beta$ (95%CI)** | **P.adjusted** | **Nnot0** | **Pathway** | **Pathway description** | **Effect $\beta$ (95%CI)** | **P. adjusted** | **Nnot0** |
| Overall | PWY.6641 | Superpathway of sulfolactate degradation | -0.33 (-0.54, -0.12) | 0.03 | 167 | PWY.6151 | S-adenosyl-L-methionine cycle I | 0.06 (0.03, 0.10) | 0.02 | 1105 |
| | HISDEG.PWY | L-histidine degradation I | -0.21 (-0.35, -0.06) | 0.06 | 1249 | PWY.7237 | Myo-, chiro- and scillo-inositol degradation | -0.27 (-0.41, -0.12) | 0.02 | 1105 |
| | SO4ASSIM.PWY | Sulfate reduction I (assimilatory) | -0.22 (-0.38, -0.06) | 0.07 | 1248 | COBALSYN.PWY | Adenosylcobalamin salvage from cobinamide I | 0.04 (0.02, 0.07) | 0.06 | 1105 |
| | PYRIDOXSYN.PWY | Pyridoxal 5'-phosphate biosynthesis I | -0.32 (-0.55, -0.08) | 0.07 | 1242 | COA.PWY | Coenzyme A biosynthesis I | 0.03 (0.01, 0.04) | 0.06 | 1105 |
| | PWY.5910 | Superpathway of geranylgeranyldiphosphate biosynthesis I (via mevalonate) | 0.57 (0.15, 1.00) | 0.07 | 1076 | PWY.5913 | TCA cycle VI (obligate autotrophs) | -0.16 (-0.27, -0.06) | 0.07 | 1105 |
| | PRPP.PWY | Superpathway of histidine, purine, and pyrimidine biosynthesis | -0.25 (-0.44, -0.06) | 0.08 | 1232 | NONMEVIPP.PWY | Methylerythritol phosphate pathway I | 0.02 (0.01, 0.04) | 0.08 | 1105 |
| | PWY0.1261 | Anhydromuropeptides recycling | -0.27 (-0.49, -0.06) | 0.09 | 1229 | PWY.7560 | Methylerythritol phosphate pathway II | 0.02 (0.01, 0.04) | 0.08 | 1105 |
| | PWY.6728 | Methylaspartate cycle | -0.44 (-0.79, -0.09) | 0.09 | 354 | P125.PWY | Superpathway of (R,R)-butanediol biosynthesis | -0.72 (-1.19, -0.25) | 0.08 | 877 |
| | PWY.6572 | Chondroitin sulfate degradation I (bacterial) | -0.43 (-0.77, -0.09) | 0.09 | 1200 | PWY.6386 | UDP-N-acetylmuramoyl-pentapeptide biosynthesis II (lysine-containing) | 0.03 (0.01, 0.04) | 0.08 | 1105 |

| | | | | | | | | | | |
|---|---|---|---|---|---|---|---|---|---|---|
| | | | | | | PWY.6385 | Peptidoglycan biosynthesis III (mycobacteria) | 0.02 (0.01, 0.04) | 0.08 | 1105 |
| | | | | | | PWY.7208 | Superpathway of pyrimidine nucleobases salvage | 0.03 (0.01, 0.04) | 0.08 | 1105 |
| | | | | | | PWY.7219 | Adenosine ribonucleotides de novo biosynthesis | 0.02 (0.01, 0.04) | 0.09 | 1105 |
| | | | | | | PWY.7229 | Superpathway of adenosine nucleotides de novo biosynthesis I | 0.02 (0.01, 0.04) | 0.09 | 1105 |
| | | | | | | PWY.6269 | Adenosylcobalamin salvage from cobinamide II | 0.04 (0.01, 0.07) | 0.10 | 1105 |
| Male | PWY.6641 | Superpathway of sulfolactate degradation | -0.27 (-0.44, -0.10) | 0.05 | 66 | No pathway was identified at P.adjusted<0.1 | | | | |
| Female | PWY.5910 | Superpathway of geranylgeranyldiphosphate biosynthesis I (via mevalonate) | 0.52 (0.20, 0.85) | 0.04 | 636 | No pathway was identified at P.adjusted<0.1 | | | | |
| | AST.PWY | L-arginine degradation II (AST pathway) | -0.67 (-1.10, -0.25) | 0.05 | 517 | | | | | |
| | PWY.5005 | Biotin biosynthesis II | 0.42 (0.13, 0.71) | 0.08 | 664 | | | | | |
| | PWY.5028 | L-histidine degradation II | -0.33 (-0.56, -0.10) | 0.10 | 107 | | | | | |

**TableS5B. Pathways associated with femoral neck bone mineral density at P.adjusted<0.1 within each study cohort; overall and sex-stratified.**

| | Rotterdam Study | | | | | Framingham Study |
|---|---|---|---|---|---|---|
| | **Pathway** | **Pathway description** | **Effect $\beta$ (95%CI)** | **P.adjusted** | **Nnot0** | |
| Overall | PWY.6353 | Purine nucleotides degradation II (aerobic) | 0.05 (0.02, 0.08) | 0.05 | 1249 | No pathway was identified at P.adjusted<0.1 |
| | PWY.7198 | Pyrimidine deoxyribonucleotides de novo biosynthesis IV | 0.07 (0.02, 0.12) | 0.05 | 1242 | |
| | PWY.5265 | Peptidoglycan biosynthesis II (staphylococci) | -0.39 (-0.66, -0.12) | 0.05 | 638 | |
| Male | No pathway was identified at P.adjusted<0.1 | | | | | No pathway was identified at P.adjusted<0.1 |
| Female | No pathway was identified at P.adjusted<0.1 | | | | | No pathway was identified at P.adjusted<0.1 |

**TableS5C. Pathways associated with trabecular bone score at P.adjusted<0.1 within each study cohort; overall and sex-stratified.**

| | Rotterdam Study | | | | | Framingham Study | | | | |
|---|---|---|---|---|---|---|---|---|---|---|
| | **Pathway** | **Pathway description** | **Effect $\beta$ (95%CI)** | **P.adjusted** | **Nnot0** | **Pathway** | **Pathway description** | **Effect $\beta$ (95%CI)** | **P. adjusted** | **Nnot0** |
| Overall | PWY.7198 | Pyrimidine deoxyribonucleotides de novo biosynthesis IV | 0.08 (0.04, 0.13) | 8.05e-03 | 1242 | METH.ACETATE.PWY | Methanogenesis from acetate | -0.18 (-0.28, -0.07) | 0.02 | 1008 |
| | PWY.7013 | L-1,2-propanediol degradation | 0.17 (0.05, 0.28) | 0.05 | 1169 | DTDPRHAMSYN.PWY | dTDP-L-rhamnose biosynthesis I | -0.02 (-0.03, -0.01) | 0.03 | 1011 |
| | | | | | | POLYAMSYN.PWY | Superpathway of polyamine biosynthesis I | 0.10 (0.03, 0.17) | 0.09 | 1011 |
| | | | | | | ARG.POLYAMINE.SYN | Superpathway of arginine and polyamine biosynthesis | 0.09 (0.02, 0.15) | 0.10 | 1011 |
| Male | PWY.7198 | Pyrimidine deoxyribonucleotides de novo biosynthesis IV | 0.10 (0.03, 0.17) | 0.09 | 518 | No pathway was identified at P.adjusted<0.1 | | | | |
| Female | No pathway was identified at P.adjusted<0.1 | | | | | No pathway was identified at P.adjusted<0.1 | | | | |

**Table S6. Association between appendicular lean mass-associated genera and maximum handgrip strength in Rotterdam Study analyzed by multivariable linear model. ***

| | **Taxa** | **Beta (95% CI)** | **P**** |
|---|---|---|---|
| Overall | Oscillibacter | -0.21 (-0.42, 9.30e-03) | 0.06 |
| | Anaerotruncus | 0.17 (-0.55, 0.21) | 0.38 |
| | Eisenbergiella | -0.37 (-0.65, -0.08) | 0.01 |
| | Agathobacter | 0.12 (-0.09, 0.33) | 0.27 |
| Female | Anaerotruncus | -0.41 (-0.75, -0.06) | 0.02 |
| | Hungatella | -0.27 (-0.55, 6.08e-03) | 0.06 |
| | DTU089 | -0.15 (-0.38, 0.08) | 0.20 |

*Adjusted covariates include age, sex, height, body fat percent, smoking (yes/no), T2D (yes/no), diet quality score, use of PPI (yes/no), use of metformin (yes/no), use of anti_lipids reagents (yes/no), sequencing batch, DNA isolation batch, time in mail, and sampling season. Taxa abundance was centered-log-ratio (CLR) transformed.

**P<0.05 is considered as significant.

**Supplementary Figure**

**Figure S1. Flowchart for sample selection in Rotterdam Study (A) and Framingham Study (B).**
Abbreviations: RS, Rotterdam Study; FHS, Framingham Heart Study.
